# Safety, Feasibility, and Utility of Digital Mobile Six-Minute Walk Testing in Pulmonary Arterial Hypertension: The DynAMITE Study

**DOI:** 10.1101/2024.08.08.24311687

**Authors:** Narayan Schütz, Vlad Glinskii, Ryan Anderson, Patricia Del Rosario, Haley Hedlin, Justin Lee, John Hess, Steve Van Wormer, Alejandra Lopez, Steven G Hershman, Vinicio De Jesus Perez, Roham T. Zamanian

## Abstract

**Rationale:** Pulmonary arterial hypertension (PAH) is a life-threatening progressive cardiopulmonary disease associated with high morbidity and mortality. Changes in the six-minute walk test (6MWT) provide prognostic information and help guide treatment decisions for PAH. However, since 6MWT requires in-clinic visits, clinical interventions to address disease progression may be delayed. Wearable technologies could reduce this delay by allowing the performance of 6MWT in the community and delivering data to clinicians remotely.

**Objectives:** To perform a pilot study to determine the safety and feasibility of performing 6MWT in PAH outpatients using a wearable app-based tool.

**Methods:** PAH patients recruited at Stanford University were provided an Apple Watch with an app to perform daily, self-administered 6MWT over 12 weeks. Bland-Altman plots and correlations were used to assess the agreement and reliability of in-clinic vs. app-based 6MWT data at the beginning and end of the 12-week trial.

**Measurements and Main Results:** From 55 PAH participants, we collected 3,139 app-recorded walks during 979.7 patient-weeks of exposure. On average, participants performed 3±2.3 weekly walks. No serious adverse events were reported. App-derived walk distance was highly correlated (*r* ≥ 0.9) to the baseline in-clinic 6MWD and showed excellent reliability (ICC=0.9). Correlation and agreement were significantly lower at the 12-week follow-up visit. App-derived metrics beyond 6MWD showed promising associations with disease status.

**Conclusions:** App-based outpatient 6MWT is feasible, safe, reasonably accurate, likely clinically relevant, and reliable in PAH patients but long-term measurement stability may be a concern. App-derived digital measures beyond distance show promise for future applications.

## INTRODUCTION

Pulmonary arterial hypertension (PAH) is a chronic cardiopulmonary disease associated with high morbidity and mortality rates despite therapy (1, 2). The six-minute walk test (6MWT) provides data on the functional capacity (3) of patients, can help identify patients at high risk of mortality who would benefit from therapeutic adjustments (4), and has been used as a primary endpoint for pivotal PAH trials. However, the 6MWT is usually performed in-clinic during routine follow-up visits and requires the supervision of trained staff to collect the data and monitor the patient’s clinical condition (5). Given that 6MWT can only be performed at clinic visits, changes in functional status can be missed and delay interventions that could reduce morbidity and mortality (6).

Recent studies have shown the feasibility of remote 6MWTs (7–9). The advantage of remote 6MWT is that the test is self-administered and is performed in the outpatient setting, providing real-time data to the healthcare provider between clinic visits. Studies have tested the feasibility of restricted and unrestricted walk approaches for remote 6MWT. The restricted walk approach mimics the in-clinic gold standard with a predefined and pre-measured course (8, 9). The unrestricted walk approach allows the patient to perform the 6MWT in the community without restrictions on location, course, or walk style (7). Although restricted walks seem more accurate, unrestricted walks have demonstrated strong correlations to the in-clinic gold standard and can be conducted with higher frequencies (7–9). However, none of these studies used a wrist-based wearable.

The goal of this longitudinal, real-world pilot study was to examine the feasibility, safety, and validity of conducting remote 6MWTs in PAH patients using unrestricted walks using a wrist-based wearable. As exploratory objective, we aimed to evaluate additional wearable derived measures for their potential clinical utility. For this study, an Apple Watch compatible app was used to capture daily 6MWT data over three months. We hypothesized that an app-based remote 6MWT would be safe to implement, demonstrate strong agreement with in-clinic 6MWT, and provide reliable data on changes in the functional capacity of patients between visits.

## METHODS

### Study Design and Participants

We designed a prospective proof of concept cohort study of PAH patients to test the safety, feasibility, and utility of remote 6MWTs. Due to COVID-19 restrictions, we modified the initial procedures to allow for remote consenting and study conduct. The study was registered (NCT03893500) and approved by the Stanford IRB. PAH patients at Stanford University Medical Center, aged 18-70 years, with WHO Group I PAH confirmed by right heart catheterization, and who owned an Apple iPhone were approached to participate. Patients stable on background PAH therapy with New York Heart Association (NYHA) functional class I-III were included. See online supplement for full inclusion and exclusion criteria.

### Study Procedures

The non-profit PHaware developed the Walk.Talk.Track™ (WTT) watch app for remote 6MWTs (https://www.phaware.global/walktalktrack). The app collected self-reported dyspnea and fatigue levels before and after a 6MWT and recorded walk data (**Figure 1, <u>Video 1</u>, <u>Video 2</u>**). Participants were provided an Apple Watch for the study and trained on the WTT app. During the initial clinic visit, demographic and clinical data were collected, and supervised ATS-guided indoor and outdoor 6MWTs (6MWT_ATS_) were conducted simultaneously with the app 6MWT. Over the next 3-6 months, participants performed daily remote 6MWTs, while following their routine clinical care plans. Zoom or telephone calls addressed questions and technical issues. Participants were re-evaluated during their next clinic appointment to gather data, collect the watch, and perform a follow-up indoor and outdoor 6MWT_ATS_.

**Figure 1.**
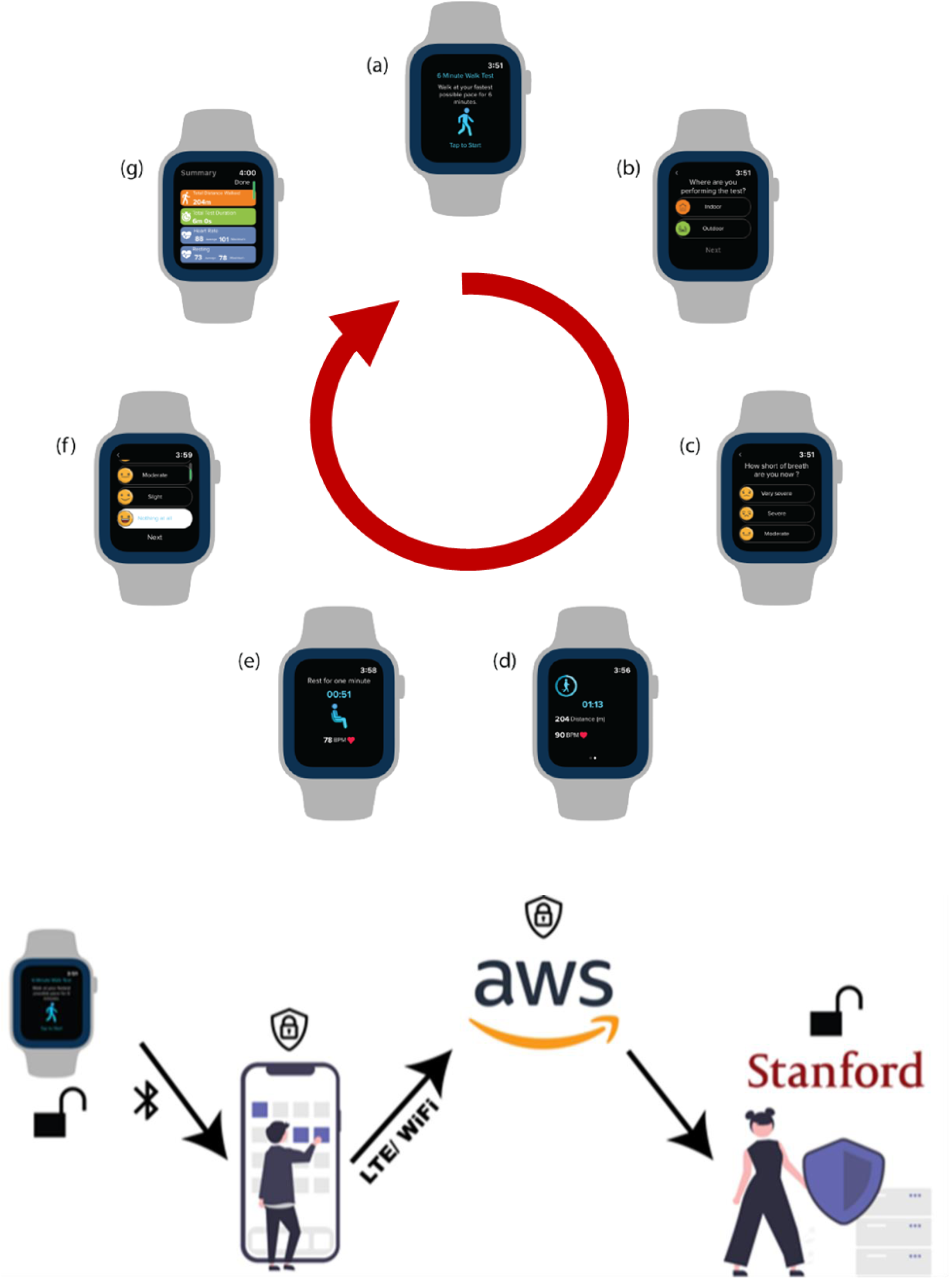
Screenshots of the Walk.Talk.Track (WTT) App interface throughout the in-community 6MWT cycle. (a) Starting screen; (b) Selection of indoor vs outdoor test; (c) Distance and heart rate during the 6-minute walk test; (e) One minute rest period; (f) Repeat two-question survey regarding the post-walk level of dyspnea and level of fatigue; (g) Summary of the walk data screen. The lower panel depicts encryption and the flow of data.

### WTT App Derived Digital Measures

We harvested 14 different variables (**Table E3**) from the WTT app, the primary two being 6MWD measures: one based on Apple’s algorithms (WTT 6MWD_R_) and another calibrated with in-clinic data (WTT 6MWD_C_). Remaining variables such as cardiac effort (9) and chronotropic index were based on heart rate and raw accelerometry.

### Statistical Analysis

Please refer to online supplement for full details.

#### Safety and Compliance

Safety was assessed based on the number and type of reported adverse events. Compliance was evaluated by calculating the number of weekly walks, with week one defined as the week of the first WTT walk. To test for a relationship between disease severity and compliance, we calculated Spearman’s *ρ* between the total number of outpatient-walks and 6MWD_ATS_.

#### WTT 6MWD Reliability & Validity

Following previous taxonomy (10, 11), we evaluated reliability by quantifying measurement errors and test-retest reliability. Measurement errors were assessed by comparing 6MWD_C/R_ against 6MWD_ATS_ during baseline and follow-up in-clinic walks and by comparing 6MWD_ATS_ with the average outpatient 6MWD_C/R_ within ±7 days of the clinic walk (adjacent-6MWD_C/R_). Systematic errors were quantified using Bland-Altman (BA) mean differences (bias), while random errors were quantified using BA 95% limits of agreement (BA95) and standard deviation (SD). Test-retest reliability was quantified using intraclass correlations (ICC). Reference values for measurement errors and ICC between same-day in-clinic walks were calculated based on indoor and outdoor 6WMTs. Long-term measurement stability(12) was quantified by comparing the error distribution of 6MWD_ATS_ and adjacent-6MWD_C/R_ between baseline and follow-up using a Mann-Whitney *U* test. Criterion validity was assessed using Pearson’s *r* between 6MWD_ATS_ and 6MWD_R/C_. Construct validity of 6MWD_C_ was assessed through discriminant, convergent, and known-groups validity. Discriminant validity was evaluated by comparing Spearman’s *ρ* between baseline 6MWD_ATS_ and adjacent-6MWD_C_ (±14-day aggregate) for cardiac output and pulmonary vascular resistance. Convergent validity was assessed similarly using NT-proBNP. Known-groups validity was assessed using NYHA FCs, with differences quantified by the Kruskal-Wallis H test. To find an optimal tradeoff between the minimum number of WTT 6MWTs per month and good reliability, we adapted a recent approach (13). The goal was to find a cut-point that minimizes the number of walks necessary while maximizing the ICC.

Hypothesis-driven tests were deemed significant at an alpha of 0.05.

### Exploratory Analyses Beyond 6MWD

WTT heart rate (HR) measures were compared with pulse oximeter during in-clinic 6MWTs. To assess measure evolution during the walk and differences between patients with varying disease status, we created time series representations of walk variables grouped by REVEAL 2.0 profiles (14) and NYHA FCs. We statistically quantified HR recovery slopes between REVEAL 2.0 risk profiles using linear mixed-effects models. To quantify the utility of HR response curve, we used a recent deep learning approach (15) that models HR response throughout the walk based on walk intensity, while automatically extracting health information. The resulting data was analyzed using linear regression.

## RESULTS

Between 02/2020 and 09/2021, 683 patients were screened. After screening through inclusion/exclusion criteria (**Figure 2**), the most common reasons for omission from the study included the inability to walk safely (N=13), the inability to perform daily walks (N=6), and concerns about the technology (N=4). Of the 62 participants consented, 7 withdrew before starting, leaving 55 patients who provided at least one walk and were considered our study population. During the study, four patients dropped out. The median cohort age was 47 [IQR 37,59] (**Table 1**). The majority of patients were women (44, 80%) diagnosed with idiopathic PAH (29%), drug and toxin-induced PAH (24%), or connective tissue disease-associated PAH (22%). No patients with NYHA class 4 symptoms were enrolled. The cohort was characterized by severe hemodynamics with a mean pulmonary arterial pressure of 46±15 mmHg and pulmonary vascular resistance of 9.1±6.5 WU. Most subjects were on either dual or triple PAH therapy (n=42, 77%). During 979.9 patient-weeks of exposure, the cohort contributed 3139 WTT walks, where 2949 walks contained valid motion (inertial measurement unit-based) data, of which 2002 also contained valid heart rate readings. Participants chose to conduct 81% (2437) walks outdoors and 19% (579) indoors (**Table E1**). A single adverse event, wrist contact dermatitis under the Apple Watch, was reported. Notably, no falls, syncopal events, or worsening of respiratory symptoms occurred during the WTT 6MWTs.

**Figure 2.**
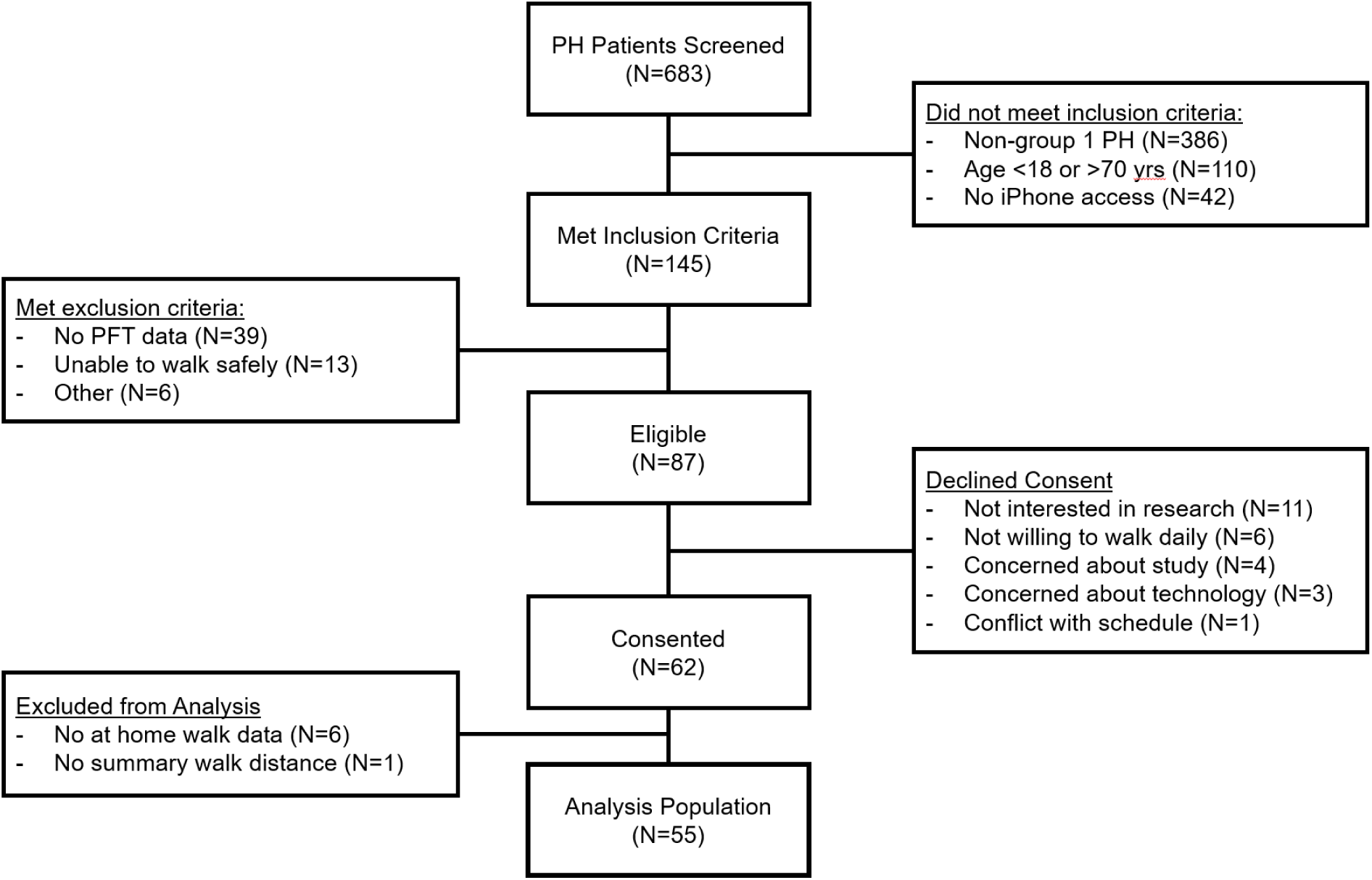
Flow diagram of the patient selection and enrollment.

**Table 1.** Patient Demographics & Clinical Characteristics.

| **Parameter** |  |
| --- | --- |
| Sex, Female, n (%) | 44 (80%) |
| Age, yrs* | 48 [37, 59] |
| BMI, kg/m<sup>2</sup> | 26.9 (5.9) |
| **NYHA Functional Class** |  |
| Class I | 15 (27) |
| Class II | 29 (53) |
| Class III | 11 (20) |
| Class IV | 0 (0) |
| **Etiologies of PAH** |  |
| iPAH | 16 (29) |
| HPAH | 4 (7.3) |
| D&T-APAH | 13 (24) |
| CTD-APAH | 12 (22) |
| Portal Hypertension | 3 (5.5) |
| CHD-APAH | 10 (18) |
| 6MWT, m | 506 (15) |
| NT-proBNP, pg/mL | 430 (116) |
| DLCO, % predicted* | 84 [66, 98] |
| Reveal 2.0 Score* | 4.0 [3.0, 6.0] |
| emPHasis-10 Score* | 12 [7, 23] |
| **Hemodynamics** |  |
| mPAP (mmHg) | 46 (15) |
| PAWP (mmHg) | 9.9 (3.5) |
| CO (L/min) | 4.70 (1.59) |
| PVR (WU) | 9.1 (6.5) |
| **Therapeutic Agents** |  |
| Endothelin Receptor Antagonist | 37 (67) |
| Phosphodiesterase Type 5 Inhibitors | 47 (85) |
| Prostacyclin | 30 (55) |
| Soluble guanylate cyclase agonist | 3 (5.5) |
| Selexipag | 2 (3.6) |
| Calcium Channel Blockers | 2 (3.6) |

Data represents mean (standard deviation), \*median [interquartile range], or number and percent where appropriate. BMI = body mass index, NYHA = New York Heart Association, PAH = pulmonary arterial hypertension, iPAH = idiopathic PAH, HPAH = heritable PAH, D&T-APAH = drug and toxin-induced PAH, CTD-APAH = connective tissue disease-associated PAH, CHD-APAH = congenital heart disease-associated PAH, 6MWT = 6-minute walk test, NT-proBNP = N-terminal pro-b-type natriuretic peptide, DLCO = diffusing capacity of the lung for carbon monoxide, mPAP = mean pulmonary artery pressure, PAWP = pulmonary artery wedge pressure, CO = cardiac output, PVR = pulmonary vascular resistance, WU = Wood units.

### Compliance

Participants were overall compliant with the 12-week protocol but an initial enthusiasm during the first two weeks was followed by a drop in daily 6MWT adherence with stabilization and recovery afterwards (**Figure 3)**. On average, participants conducted 3±2.4 walks per week (**Figure E1**), and many participants continued routine 6MWTs beyond 12 weeks (**Figure 3**). Overall, approximately 60% of participants performed at least one walk every single week throughout the entire study (**Figure E2**), and ∼20% of participants performed 6-7 walks per week.

**Figure 3.**
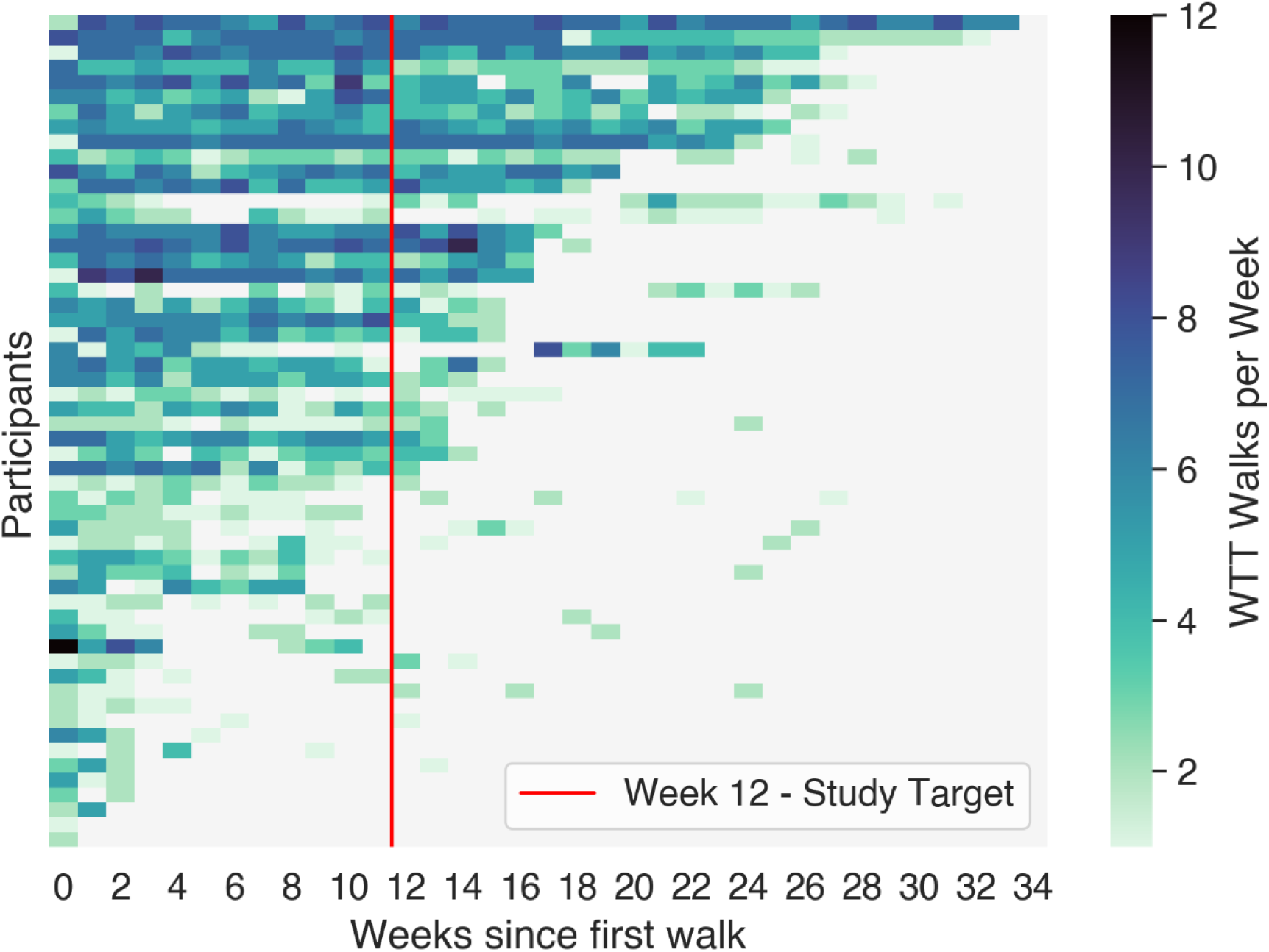
Heatmap of walks performed per week for each participant with at least one at-home WTT app walk. The study officially ended at week 12, but many participants chose to continue walking for up to 34 weeks. Some participants even opted to perform more than one at-home walk per day.

### Reliability of WTT-Derived 6-Minute Walk Distances

Comparing in-clinic gold standard 6MWD_ATS_ against WTT 6MWDs, we found mixed results (**Table 2**). WTT 6MWD_R_ (based directly on Apple’s distance estimation algorithm) showed relatively high variation in the difference between the two measures with standard deviations between 75-85 m and high BA95 (**Figure 4**); moreover, the outdoor walks showed a sizable bias of -66.16 m. Our calibration model (WTT 6MWT_C_) reduced the bias to -2.61±40.09 meters (**Figure 4**), resulting in excellent test-retest reliability (ICC=0.9, n=44) and plausible distribution of community walks (**Figure 5)**. For reference, the ICC of same-day 6MWD_ATS_ was 0.92 (at baseline with n= 33 participants). A similar pattern of almost equivalency between same-day 6MWD_ATS_ variations and variations between 6MWD_ATS_ vs adjacent-6MWD_R/C_ was observed based on SD and BA95 (**Table 2**, **Figure E10**). In terms of long-term measurement stability, both WTT 6MWDs (WTT 6MWD_C_ not reported) diverged from the in-clinic 6MWD_ATS_ over time, resulting in a larger error (bias, BA95, and SD) for the follow-up visits (**Figures E4 & E7**). Statistically, the difference in measurement error distributions between baseline and follow-up was significant (Mann-Whitney *U*=260, *P*=0.0015).

**Figure 4.**
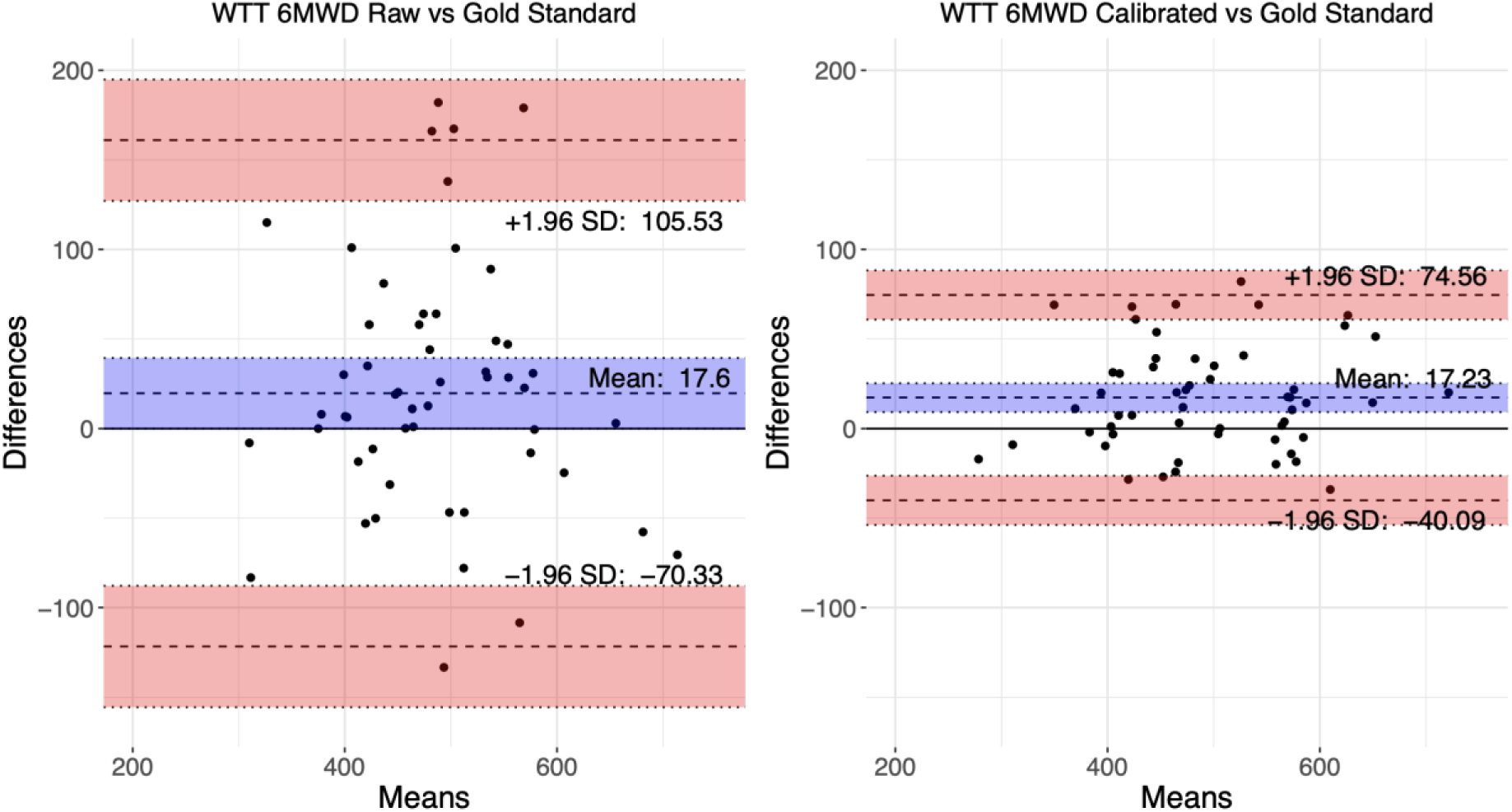
The Bland-Altman plots show the agreement between in-community WTT 6MWDs from walks conducted within seven days of the clinic visit and the actual in-clinic gold standard. The left panel presents the WTT 6MWD Raw (n=54), while the right panel highlights the WTT 6MWD Calibrated (n=54).

**Figure 5.**
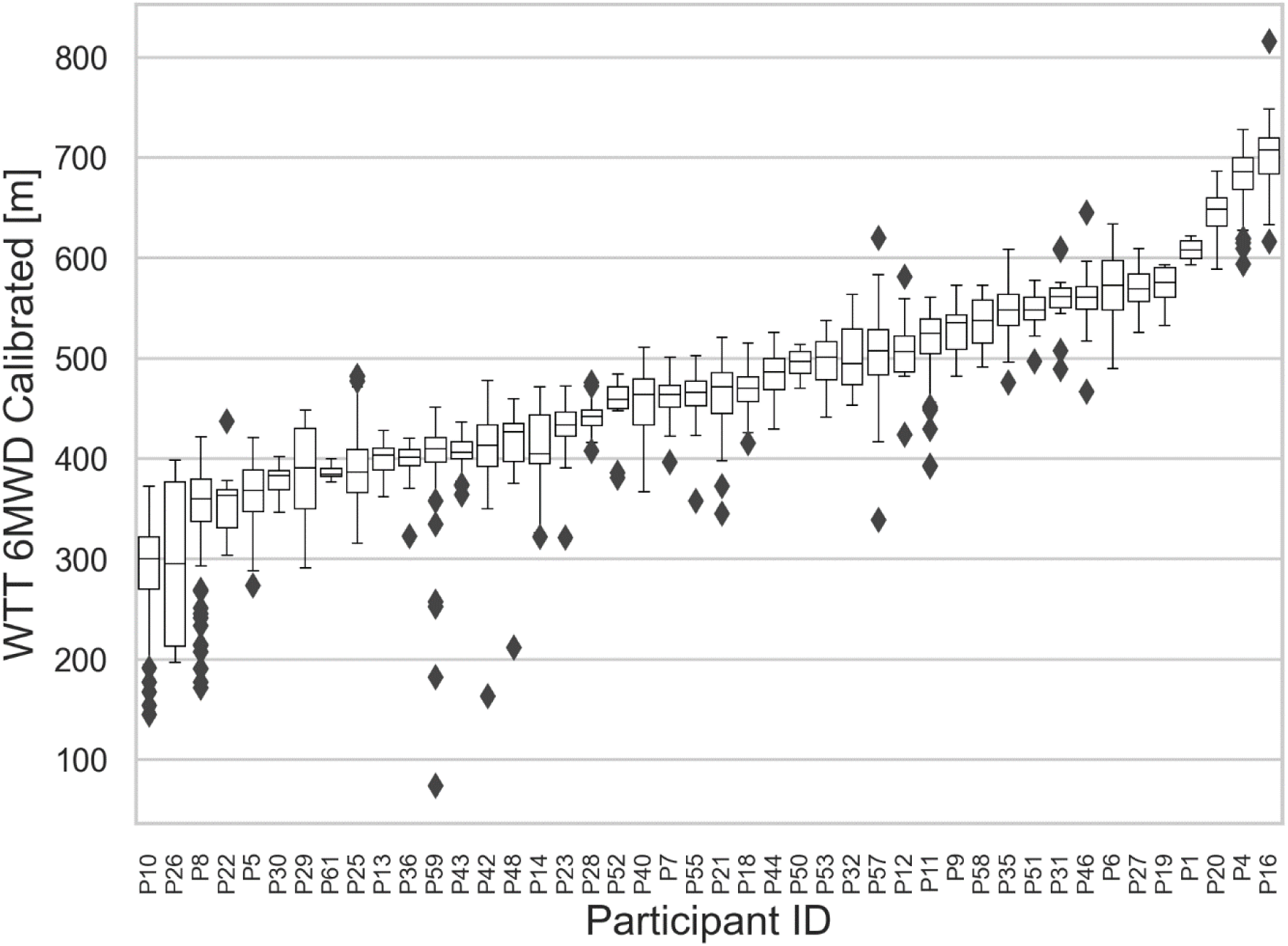
At-home WTT 6MWD Calibrated distribution for each participant with valid at-home walk data.

**Table 2.** Gold Standard vs. WTT 6MWD Agreement Comparison.

| 6MWD<br>Measurement 1 | 6MWD<br>Measurement 2 | N | Distance (m)<br>mean±SD | *r* (*P*) |
| --- | --- | --- | --- | --- |
| **In-Clinic** |  |  |  |  |
| Gold Standard<br>Indoor | WTT 6MWD<sub>R</sub><br>In-clinic Indoor | 54 | -11.30±77.33 | 0.78 (< 0.001) |
| Gold Standard<br>Outdoor | WTT 6MWD<sub>R</sub><br>In-clinic Outdoor | 48 | 62.09±87.15 | 0.39 (< 0.001) |
| Gold Standard<br>Outdoor | WTT 6MWD<sub>C</sub><br>In-clinic Outdoor | 48 | -2.61±40.09 | 0.91 (< 0.001) |
| **In-Clinic vs. Adjacent At-Home** |  |  |  |  |
| Gold Standard (all)<br>Baseline | WTT 6MWD<sub>R</sub><br>in-community | 54 | 17.6±44.86 | 0.9 (< 0.001) |
| Gold Standard (all)<br>Follow up | WTT 6MWD<sub>R</sub><br>in-community | 19 | 74.29±71.26 | 0.73 (< 0.001) |
| Gold Standard<br>Baseline | WTT 6MWD<sub>C</sub><br>in-community | 54 | 17.23±29.25 | 0.95 (< 0.001) |
| Gold Standard<br>Follow up | WTT 6MWD<sub>C</sub><br>in-community | 19 | 57.84±64.43 | 0.76 (< 0.001) |
| **In-Clinic vs. In-Clinic** |  |  |  |  |
| Gold Standard<br>Indoor | Gold Standard<br>Outdoor | 43 | 23.30±24.0 | 0.94 (<0.001) |

6MWT<sub>R</sub> = 6MWT Raw based on Apple's proprietary algorithm, 6MWT<sub>C</sub> = 6MWT Calibrated

### Validity and Reliability of WTT-Derived 6-Minute Walk Distances

Concerning criterion validity, mean aggregated at-home adjacent-6MWD_R_ and adjacent-6MWD_C_ were strongly correlated with the 6MWD_ATS_ at baseline (Pearson’s *r*=0.9 and *r*=0.95, respectively), which is in line with the correlation between the two in-clinic walks at the baseline visit (*r* = 0.94) (**Table 2)**. Regarding construct validity, associations of baseline 6MWD_ATS_ and 14-day adjacent WTT 6MWD_C_ with established clinical variables showed congruent results for the two 6MWDs in terms of convergent, discriminant and known-groups validity (**Table 3**). We found that reliability increases considerably from an ICC of 0.85 at one walk per month to 0.93 at 14 walks per month. An optimal tradeoff between number of walks and reliability was found at 4 walks per month corresponding to an ICC of 0.91 (**Figure E9**).

**Table 3.** Construct Validity of ±14 Day Mean Aggregated adjacent-6MWD_C_ around Baseline.

| Measurement | Stat 6MWD<sub>ATS</sub> (*P*) | Stat 6MWD<sub>c</sub> (*P*) | Statistic |
| --- | --- | --- | --- |
| Cardiac Output*<br>(L/min) | 0.00 (0.99) | -0.05 (0.976) | Spearman's $\rho$ |
| Pulmonary Vascular<br>Resistance<br>(dyn/s/cm<sup>-5</sup>) | -0.17 (0.35) | -0.08 (0.67) | Spearman's $\rho$ |
| NT-proBNP (pg/mL) | -0.4 (0.02) | -0.42 (0.01) | Spearman's $\rho$ |
| NYHA Class | 9.66 (0.008) | 6.43 (0.04) | Kruskal-Wallis *H* |

\*Spearman's $\rho$ for all variables except, Kruskal-Wallis test for NYHA functional class.

### Beyond Six-Minute Walk Distance

The potential novelty of WTT technology is the ability to measure or derive other variables than the walk distance. Summary statistics for all 14 WTT measures are presented in **Table 4**. Group differences that one could expect between different NYHA FCs can be observed. Similar differences can also be observed between walk variable trajectories. For instance, WTT Mean Amplitude Deviation (a measure of exercise intensity) increases over walk-time for NYHA class 1 participants but appears constant or slightly decreasing for class 3 participants (**Figure 6**). Similar patterns were evident when grouping by post-walk dyspnea ratings (**Figure E8**). Beyond NYHA, clearly discernible differences are visible when grouping participants by REVEAL 2.0 risk profiles. Moreover, we observed a statistically significant difference between post-walk recovery differences amongst different risk groups, where low-risk participants recover faster after 1-minute post-walk (slope = -0.28 beats/s CI±0.006) than moderate-risk participants (slope = -0.20 beats/s CI±0.012), and those recover faster than high-risk participants (slope = -0.03 beats/s CI±0.021) (*P*=<0.001) (**Figure 7).** Heart rate measurements by the WTT app further showed strong correlations to the pulse oximeter-derived with Pearson’s *r* of 0.82 (n=33) for Peak HR and *r* of 0.9 (n=33) for post-walk HR (**Figure E5**). Finally, the automatically extracted variables from heart rate response curves using a deep learning approach explained 50.8% (R^2^=0.508, n=36) of the variance when regressing REVEAL 2.0 scores (**Figure E11)**, while regressing NYHA FC accounted for 48.6% (R^2^=0.486, n=38) of the variance.

**Figure 6.**
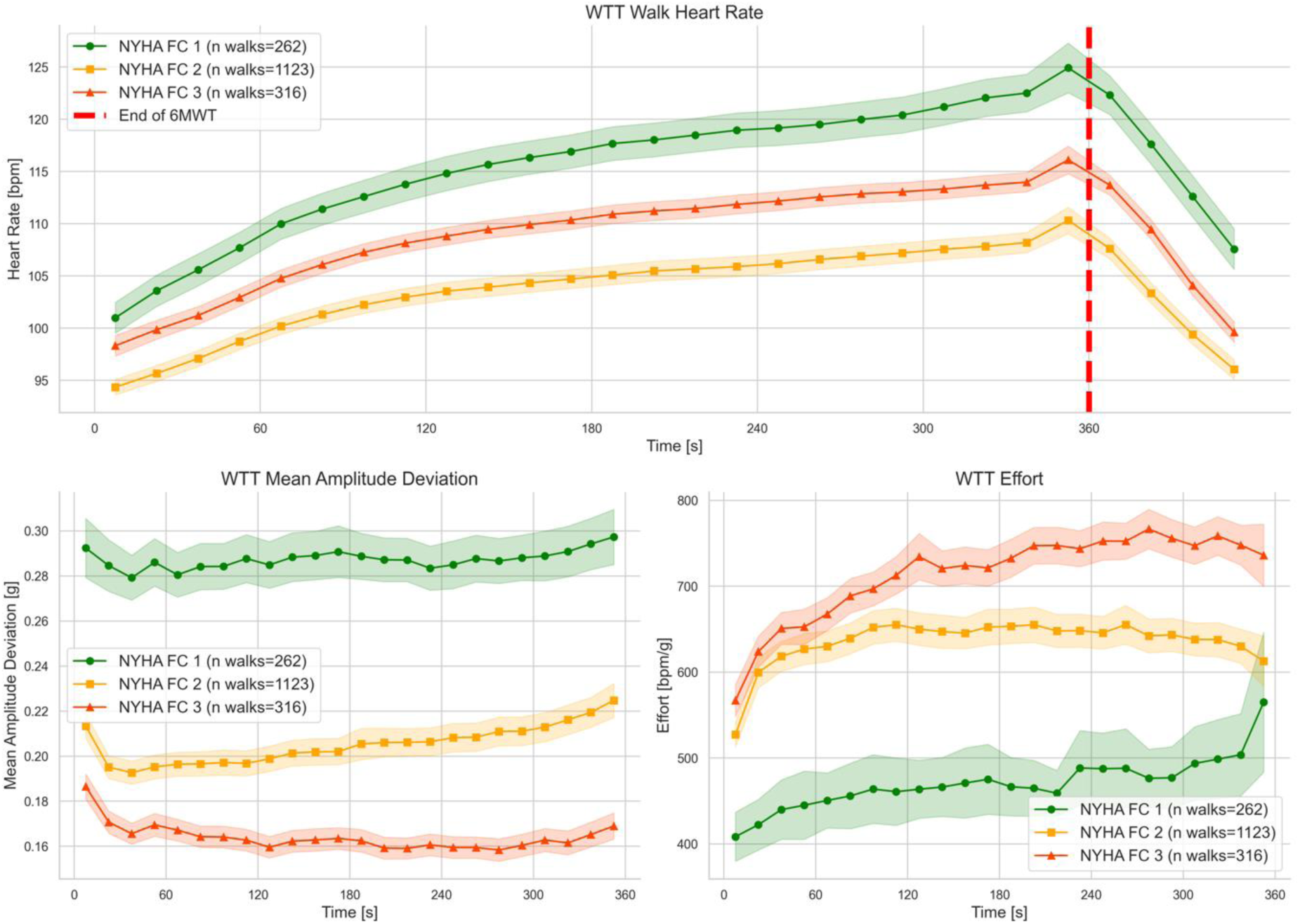
Multiple WTT variables over the 6 minutes. All walks are aggregated across time and grouped by NYHA Functional Class. A) WTT Heart Rate refers to the Apple provided heart rate throughout and 1min after the walk; B) WTT Mean Amplitude Deviation is a measure of physical activity intensity based on raw accelerometer data throughout the walk; C) WTT Effort relates to continuous pulse estimates to continuous physical using a ratio, where higher values indicate greater effort. Besides revealing apparent differences between functional classes, the illustration also shows differences in the dynamics over time, particularly in the case of physical activity intensity, where FC 1 and 2 show an increase in intensity over time while FC 3 participants seem to decrease.

**Figure 7.**
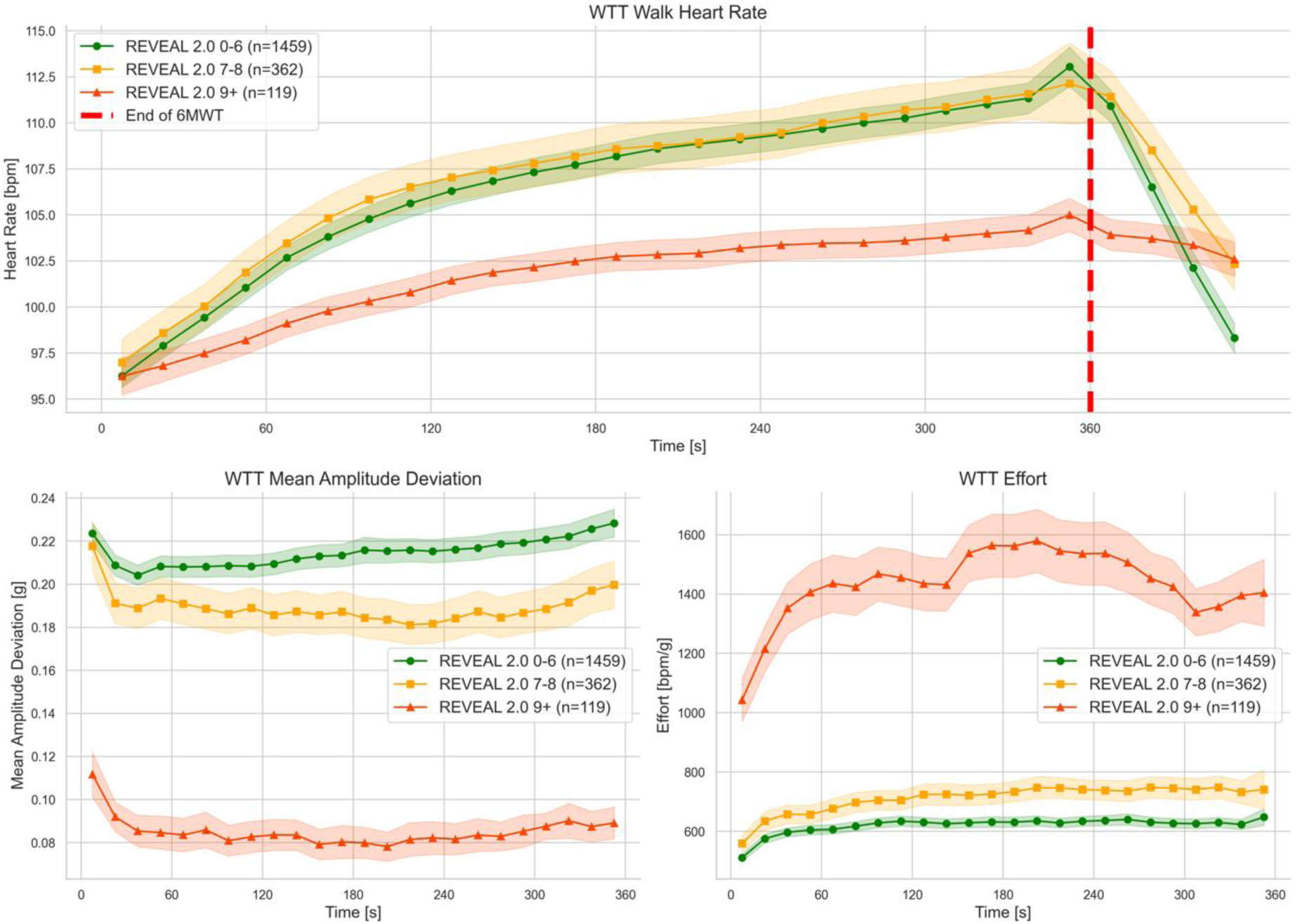
Multiple time binned WTT variables across different REVEAL 2.0 Risk states. This highlights how, on average, walks from patients with a specific risk category, evolved over the 6-minute walk duration. A) WTT Heart Rate refers to the Apple provided heart rate throughout and 1min after the walk; B) WTT Mean Amplitude Deviation is a measure of physical activity intensity based on raw accelerometer data throughout the walk; C) WTT Effort relates to continuous pulse estimates to continuous physical using a ratio, where higher values indicate greater effort.

**Table 4.** Summary Statistics of WTT Variables.

| WTT Variable | ICC | Total | NYHA FC 1 | NYHA FC 2 | NYHA FC 3 |
| --- | --- | --- | --- | --- | --- |
| WTT 6MWD Raw | 0.85 | 472 (43) | 500 (44) | 467 (44) | 419 (39) |
| WTT 6MWD Calibrated | 0.9 | 473 (32) | 507 (29) | 467 (32) | 405 (37) |
| WTT Steps | 0.8 | 645 (42) | 656 (39) | 649 (43) | 595 (50) |
| WTT Total MAD | 0.81 | 786 (148) | 854 (142) | 812 (164) | 468 (76) |
| WTT HR Reserve | 0.83 | 94 (8) | 98 (9) | 94 (8) | 84 (4) |
| WTT Chronotropic Index | 0.7 | 0.44 (0.09) | 0.51 (0.10) | 0.41 (0.09) | 0.39 (0.07) |
| WTT HR Recovery | 0.6 | 16 (7) | 17 (8) | 16 (7) | 11 (4) |
| WTT Peak HR | 0.78 | 119 (8) | 125 (10) | 117 (8) | 111 (5) |
| WTT HR Expenditure | 0.8 | 640 (39) | 666 (44) | 634 (39) | 611 (26) |
| WTT Cardiac Effort | 0.72 | 1.38 (0.14) | 1.35 (0.11) | 1.39 (0.15) | 1.44 (0.12) |
| WTT Pre-Walk Dyspnea | 0.61 | 0.44 (0.29) | 0.40 (0.34) | 0.40 (0.26) | 0.80 (0.40) |
| WTT Post-Walk Dyspnea | 0.7 | 1.16 (0.43) | 1.28 (0.38) | 1.00 (0.44) | 1.71 (0.51) |
| WTT Pre-Walk Tiredness | 0.62 | 0.59 (0.36) | 0.59 (0.39) | 0.54 (0.33) | 0.91 (0.47) |
| WTT Post-Walk Tiredness | 0.72 | 1.00 (0.42) | 1.10 (0.46) | 0.89 (0.40) | 1.46 (0.52) |

Data represent Mean (Standard Deviation). WTT=Walk.Talk.Track, ICC=intraclass coefficient, NYHA FC=New York Heart Association Functional Class, MAD=Mean Amplitude Deviation, HR=Heart Rate, bpm=beats per minute, m=meters

## DISCUSSION

This pilot study evaluated the feasibility, safety, and clinical data output from app-based 6MWTs performed on PAH patients in the community. The absence of a significant adverse event in 3139 community 6MWTs suggests that PAH patients can safely self-administer these tests, wherein they have the freedom to decide where and how to perform the walk. This data supports the argument that unrestricted 6MWTs can be safely self-administered by PAH patients (7–9, 16). Regarding clinical utility, results show that variables derived from community 6MWTs are highly correlated with in-clinic 6MWTs and when mean aggregated show very similar agreement, correlation and test-retest reliability as can be observed between pairs of same-day clinic 6MWTs. Notably, the study raises concerns over the long-term measurement stability of such remote walks indicated by an increasing deviation from the in-clinic estimates over time in some participants.

Our findings indicate that compliance with the daily walks mandate is variable. Some participants stopped providing daily walks early in the study, while others continued to use the WTT app daily even after the study was concluded. No significant correlation was found between disease severity (based on in-clinic 6MWD_ATS_) and test compliance, indicating that disease severity may not have been a limiting factor. However, despite the mixed compliance, most patients demonstrated greater engagement with the outpatient 6MWT compared to a similar study in PAH patients, where patients struggled with monthly targets (7). Several factors could account for this, including emphasizing a patient-centric and user-friendly strategy that did not require additional devices or specific smartphone handling.

In terms of reliability and validity, our results are promising. We found a systematic underestimation of 6MWD of 17m when comparing 6MWD_ATS_ with adjacent-6MWD_R/C_ at baseline. Using our proposed calibration approach, we could not reduce this bias; however, we could meaningfully reduce random measurement error, with a reduction in SD from 64m to 29m going from uncalibrated to calibrated adjacent-6MWD and similar reductions in Bland-Altman 95% limits of agreement (both in-clinic and in-community). Similarly, test-retest reliability was good (ICC=0.85) and excellent (ICC=0.9) for both the uncalibrated and calibrated 6MWD, respectively. Taken together, these results suggest that WTT derived 6MWD provides a reliable measurement of in-clinic 6MWD in the unconstrained community setting. While individual 6MWD_C/R_ values are similar, they are not close enough to the in-clinic ground truth to warrant interpretation as a drop-in replacement, considering that a minimal clinically relevant change is 33m (17). However, when outpatient walks are aggregated over a time-interval of 14-30 days, differences between community and adjacent in-clinic tests were almost equivalent to the differences observed between same-day in-clinic walks (**Table 2**). Looking at different construct validity measurement properties further strengths these results, highlighting high correlations (criterion validity) of 6MWD_R/C_ to the gold standard (>0.9), at least at baseline (**Table 3**). Moreover, 6MWD_C_ exhibits an expected (18, 19) correlation with NT-proBNP (comparable to 6MWD_ATS_), a lack of significant correlations with pulmonary vascular resistance and cardiac output (comparable to that of 6MWD_ATS_), as well as expected group-differences between NYHA FCs (**Table 3)**.

Reliability and validity data was promising at baseline, but we observed a marked increase in both systematic and random errors at follow-up (**Figure E4**). This increase is not only statistically significant but also qualitatively visible when looking at the curves of individual participants. While not present in all participants, this effect has not been previously reported but could be highly relevant for future digital outcome assessments that require frequent patient effort, essentially highlighting the importance of validating such instruments longitudinally. One explanation could be that some patients anchor their effort on previous walks while gradually reducing relative effort over time. This notion of reduced effort is supported qualitatively by decreasing walk HR and decreasing self-rated WTT Post-Walk Dyspnea ratings across individual case studies (**Figure E7**). These indicators could thus help identify decreasing effort but more data would be needed to come to conclusions. While it is unclear how to address this phenomenon, a potential way could involve motivating patients to surpass their previous walk performance, or reducing administration frequency. A recent study in pulmonary hypertension did not report a drop in agreement but only required two at-home tests between clinic visits (16). This gives some credibility to the idea of reduced frequency. Here it is noteworthy that our results suggest that excellent reliability can be achieved with only four walks per month. Another avenue may be introducing the notion of cardiac effort instead of 6MWD, which divides the total heartbeats by the 6MWD (9). However, in our case, this led to reduced test-retest reliability, resulting in an ICC of only 0.72, possibly caused by low-quality heart rate readings.

Beyond traditional 6MWD, digitally recorded 6MWTs also promise to record additional measures during a 6MWT, including continuous metrics throughout the walk (9). These include accelerometry derived values, such as WTT Total Mean Amplitude Deviation (MAD), a measure of activity intensity (20), or continuous pulse estimation that could allow monitoring of previously ignored characteristics (21, 22). In this direction we found how averaged WTT MAD curves differed across NYHA classes (**Figure 6**) and REVEAL 2.0 risk profiles (**Figure 7**). While clinical interpretation remains to be established, this additional information could provide novel insights complementing the traditional 6MWD without additional effort to personnel. The same goes for app-derived patient-reported outcomes, such as the self-reported dyspnea and tiredness ratings in this study, the former of which we found associated with walk variables (**Figure E8**) and which could help characterize patient effort or perceived changes in exercise tolerance. In this vein, our results indicate that these new continuous data streams could hold significant potential when combined with advanced artificial intelligence approaches. Preliminary findings suggest that the relationship between walk intensity and heart rate response provides valuable insights into cardiorespiratory health and fitness. Extracted data from these dynamics could explain approximately 50% of the variation in both NYHA functional classes and REVEAL 2.0 risk scores. While experimental, these results suggest that heart rate response dynamics could hold relevant health information in PAH patients and may warrant further investigation.

A major limitation of this study is that no NYHA class 4 patients were included as it was too risky before first establishing safety in class 1-3 patients. Another limitation is the significantly lower number of reliable heart rate readings (only about 2000 out of 3000 walks). Other studies have also found that measuring heart rate using photoplethysmography during 6MWTs is not as accurate even when based on a medical device (7, 9), making motion artifacts a plausible explanation and raising the question of whether a wrist-worn device is the right choice when it comes to heart rate during 6MWTs. Moreover, this is a single-center study, which may limit generalization to different sites (for instance, due to different climate conditions). Lastly, it is relevant to note that while technical issues with the WTT app were minimal, and patients were told to reach out in case of any issues, it was impossible to distinguish app failure from failure to comply with the study protocol. As such, compliance may be underestimated.

In conclusion, this pilot study confirms that unrestricted community-based 6MWTs for PAH patients are both feasible and safe, with no serious adverse events and reliable app-based 6MWD. Although single app-based walks show more variation, averaging measurements over 14-30 days produces results comparable to in-clinic tests. The study also found good construct validity for the app-based 6MWD across various measurement properties, with weekly community 6MWTs providing excellent test-retest reliability for many PAH patients. However, challenges include long-term measurement stability, with increasing deviation from in-clinic 6MWD estimates over time and suboptimal heart rate quality. Despite these challenges, the digital 6MWT offers additional insights without extra effort from patients or staff, including heart rate variables, continuous walk data, and the potential to use advanced artificial intelligence to extract more complex health information. Future research should focus on NYHA functional class 4 patients, continuous walk variables, heart rate dynamics, improving heart rate quality, and maintaining patient motivation over time.

## Data Availability

All data produced in the present study may be available upon reasonable request to the authors.

https://www.phaware.global/walktalktrack

https://vimeo.com/876424176/faff471270

https://vimeo.com/872681227/ba027be5ed

## ACKNOWLEDGEMENTS

We want to thank all the patients who participated in our study.

## Author Contributions

VG, RA, JH, SVW, VDJP, and RTZ contributed to the study design. VG, RA, PD, AS, and AL contributed to data collection. NS, VG, and RA conducted data analysis. All authors contributed to manuscript writing and editing.

