## Supplement for "Safety, Feasibility, and Utility of Digital Mobile Six-Minute Walk Testing in Pulmonary Arterial Hypertension: The DynAMITE Study"

**ONLINE DATA SUPPLEMENT**

**Supplementary Methods**

**Study Procedure Further Details**

Patients who were impacted by the COVID-19 pandemic and could not attend the first appointment were remotely consented, sent an Apple Watch, and conducted a remote Zoom teaching session with the clinical research coordinator.

**Participant Inclusion Criteria**

1. Age ≥ 18 and < 75 years

2. Diagnosis of WHO Group I Pulmonary Arterial Hypertension (PAH) (Idiopathic (I)PAH, Heritable PAH (including Hereditary Hemorrhagic Telangiectasia), Associated (A)PAH (including collagen vascular disorders, drugs + toxins exposure, congenital heart disease, and portopulmonary disease).

3. Any Previous Right Heart Catheterization that documented:

a. Mean PAP ≥ 25 mmHg.

b. Pulmonary capillary wedge pressure ≤ 15 mmHg.

c. Pulmonary Vascular Resistance ≥ 3.0 Wood units or 240 dynes/sec/cm5

4. All NYHA/WHO functional classes.

5. Be an owner of an iPhone

**Participant Exclusion Criteria**

1. WHO Group II – V Pulmonary Hypertension

2. TLC < 60% predicted; if TLC b/w 60 and 70% predicted, high-resolution computed tomography must be available to exclude significant interstitial lung disease

3. FEV1 / FVC < 65% and FEV1 < 60% predicted

4. Inability to perform a 6-minute walk test (6MWT)

5. Significant left-sided heart disease (based on screening Echocardiogram):

    a. Significant aortic or mitral valve disease

    b. Diastolic dysfunction > Grade II

    c. LV systolic function < 40%

    d. Pericardial constriction

    e. Restrictive cardiomyopathy

    f. Significant coronary disease with demonstrable ischemia.

6. Current atrial arrhythmias not under optimal control.

7. Uncontrolled systemic hypertension: SBP > 160 mmHg or DBP > 100mmHg

8. Severe hypotension: SBP < 80 mmHg

9. Psychiatric, addictive, or other disorder that compromises participant's ability to provide informed consent, to follow study protocol, and adhere to treatment instructions

10. Co-morbid conditions that would impair a participant's exercise performance and ability to assess WHO functional class, including but not limited to chronic low-back pain or peripheral musculoskeletal problems

**Data Flow Details**

Upon completion of the walk, the Apple Watch part of the WTT application synced the walk data to the iPhone pendant of the app, where it was locally encrypted using public-key encryption and then transmitted to AWS S3 for storage. Walk data was eventually downloaded to Stanford infrastructure certified to handle protected health information (PHI) and only eventually decrypted on-site. Only authorized Stanford personnel had access to the private key, ensuring no unauthorized third party would in any way be possible to access any WTT data.

**WTT Data Preprocessing and Data Cleaning:**

WTT walks measured by the device were filtered using two groups of exclusion criteria. The first group pertained to motion and pedometer data, while the second concerned heart rate data. To incorporate as many walks as possible, we evaluated distance agreement (detailed below) using the motion and pedometer data.

Conversely, all other analyses were conducted on walks that satisfied both sets of criteria. Furthermore, we excluded any walks in which the time reported by the app deviated by more than ±15 seconds from the 6-minute mark, irrespective of the sensor data measurements.

The walk exclusion criteria based on motion data included: (1) complete absence of motion or pedometer data; (2) reported distances of < 110m or < 100 reported steps; (3) instances where motion or pedometer data were reported for < 350 seconds; (4) cases where the total number of raw motion sensor data samples deviated by >50 from the target of 3600 samples.

Filtering by heart rate data incorporated the following criteria: (1) < 54 HR samples; (2) < 6 HR samples for any given minute; (3) instances where no post-walk resting HR data was recorded.

The actual number of walks per analysis was further influenced by the necessity of many analyses to have at least one indoor and outdoor gold standard walk (to fit the distance estimation model), to keep results as transparent as possible the involved number of participants is reported with each result.

Walks with walk identifier "*75ff08d9-1a78-44a5-afe5-7a120cc4ccab*" and "*da760287-0563-4faf-b60f-0e74889ca593*" were excluded manually, as they contained invalid walk data.

**Apple Watch**

Participants were provided with a Series 4 or 5 Apple Watch for the duration of the study.

**WTT Variable Statistics**

To evaluate the potential of app-derived measures beyond 6MWD, we calculated grand mean, average SD, and ICC across all participants and walks. We also calculated these statistics grouped by NYHA FCs.

**Detailed 6-Minute Walk Distance and Heart Rate Agreement Evaluation**

Agreements between gold standard in-clinic and WTT-based $6MWT_{ATS}$ were evaluated with Bland-Altman plots and numerically quantified using $R^2$ statistics. All participants with the necessary demographics data and concurrent in-clinic and WTT-based 6MWT data were included. Two walks were excluded due to invalid gold standard 6MWD values entered in the case report form. For the comparison of gold standard 6MWDs vs at-home WTT-derived 6MWDs, at-home walks within one week (in both directions) of the clinic visit were mean aggregated and compared. To include as many walks as possible, the average of indoor and outdoor gold-standard $6MWD_{ATS}$ was used in cases with both were available.

WTT Peak HR was compared against peak exercise and 2 min post-walk HR obtained from a medical grade oximeter, using the same concurrent data as was used to evaluate 6MWD. It is important to note the measurements from the oximeter were taken at slightly different time points. While peak heart rate with the WTT app is based on the average of the last three heart rate samples of the 6-minute walk test, the oximeter measurement was taken directly after the walk. Similarly, the WTT-based HR for resting was defined as the three last HR samples 1 min after the walk, while the oximeter-based HR was taken after 2 min of resting post-walk. For this analysis, we included all available indoor in-clinic walks that had simultaneous WTT data available.

**Heart Rate Recovery Slope Comparison**

We fit a linear mixed effects model to model the heart rate over time with random intercepts for participant and walk to account for repeated observations. We used a likelihood ratio test (between a model with the grouping variable as an interaction term and a default model without) to evaluate whether the heart rate slopes over time differed between groups.

**Leveraging Sensor Data and Machine Learning to Model Heart Rate Response**

We made use of a recently published approach that models the evolution of heart rate during exercise using ordinary differential equations (ODEs), where the parameters of the ODEs are estimated through neural networks (1). The model learns a latent representation for each participant, supposably encoding their cardiopulmonary health and fitness status. We made use of the original author's implementation (https://github.com/apple/ml-heart-rate-models). We trained the model using the publicly available *Endomondo* exercise dataset (2) and 1596 (that fulfilled pre-processing criteria: distance walked > 300m; minimum number of walks per patient >= 6; max speed between 2 and 40 km/h; at least one heart rate reading every 50s; min HR >= 40 bpm; max HR < 215 bpm, GPS data available), WTT 6MWTs from our PAH patient population. The model was trained with a *batch_size* of 256, *history_max_length* of 512, *chunk_size* of 8, and *stride* of 8 for 10 epochs. The remaining parameters were left at their default values. For each participant, 20% of the most recent walks were used as test set. We didn't include any weather information. For WTT app-derived horizontal distance data we used Apple's provided pedometer data. The 10-dimensional learned health and fitness representations for each participant were (based on the test data) then used to regress New York Heart Association functional classes (NYHA FCs) and REVEAL 2.0 scores using an ordinary least square fitted multivariate linear regression model.

**Calculating Intraclass Correlations**

All intraclass correlations (ICCs) were calculated using *lme4* in R by fitting a model of the form "*VARIABLE ~ (1|participant_id)",* where VARIABLE corresponds to the target variable that we want to calculate the ICC of. Using this model we then calculate the ratio measuring the proportion of variance that is attributable to differences between participants. Thus, $ICC = \frac{s_w^2}{s_w^2+s_b^2}$, where $s_w^2$ refers to the variance between participants and $s_b^2$ to the residual variance.

**Walks per Month to Reach Reliability**

To determine how the number of WTT 6MWTs per week influences test-retest reliability we split up the walks of all participants into 4-week non-overlapping intervals and selected those that contain at least 14 walks (thus half the target frequency of this study). To maximize the number of participants that could be included, we used the uncalibrated $6MWD_R$ (some participants only had one only one walk but we needed at least two for the calibration). This resulted in a total of 94 4-week intervals from 28 unique participants. We subsequently uniformly sampled $k \in \{Z: 1 \leq k \leq 14\}$ walks from each participant's 4-week interval, mean aggregated the resulting walk distance, and calculated the ICC. This procedure was repeated 100 times to account for the sampling process (thus selecting any subset of walks from each 4-week interval).

To determine the ideal trade-off between a high ICC and a low number of necessary walks, we aggregated the ICC across the 100 samples, yielding a more stable result. Subsequently, we (Min-Max) normalized both the sample and ICC dimension and determined the point with the minimal Euclidean distance to the left top corner (1,1). The optimal point and full curves are visualized in **Figure E9**.

**Software Used and Statistics**

General data preprocessing and visualization were conducted using Python v3.8.16, SciPy v1.9.3, NumPy v1.23.5, and Pandas v1.5.1. Bland-Altman plots, mixed effects models, and ICCs were computed with R v3.6.2 using the "blandr" v0.5.1 and lme4 v1.1-24 packages. All other employed statistical tests where calculated using the SciPy package in Python.

**Supplementary Results**

When comparing adjacent-$6MWD_R$ to the in-clinic ground truth we found that both WTT $6MWD_R$ and $6MWD_C$ estimates underestimated $6MWD_{ATS}$ by around 17m for the baseline visit and 74m for the follow-up visit, respectively (Figure E4). Visually inspecting time-series values further confirmed these results, showing that many participants' WTT 6MWDs (both calibrated and raw) diverged from the in-clinic value over time, resulting in a larger error for the follow-up visits. A case example of this behavior is shown in **Figure E7**.

**Leveraging Sensor Data and Machine Learning to Model Heart Rate Response**

Beyond the reported results in the main manuscript, we found that the mean absolute error for heart rate prediction was 7.613 bpm on the test workouts. Estimated HR max was markedly lower in PAH patients as opposed to users from the *Endomondo* dataset with 159.8 $\pm$ 7.4 bpm vs 189.8 $\pm$ 11.9 bpm, respectively. For reference, HR max based on the common formula $220 - age$ resulted in an estimate of 172 bpm within our PAH study population.

**Supplementary Tables**

**Table E1. WTT App Walk Numbers**

| Description | Number |
|---|---|
| Total In-clinic walks with WTT | 110 |
| Total community walks received | 3139 |
| Patient weeks of exposure | 979.7 |
| Total walks indoors | 579 (19.6) |
| Total walks outdoor | 2437 (82.6) |

Numbers represent totals, numbers, and percent where appropriate.
WTT = Walk.Talk.Track

**Table E2. Intraclass Correlations of WTT Variables**

| WTT Variable | ICC |
|---|---|
| WTT 6MWD Raw, m | 0.85 |
| WTT 6MWD Calibrated, m | 0.9 |
| WTT Steps, count | 0.8 |
| WTT Total MAD, g | 0.81 |
| WTT HR Reserve, BPM | 0.83 |
| WTT Chronotropic Index | 0.7 |
| WTT HR Recovery, BPM | 0.6 |
| WTT Peak HR, BPM | 0.78 |
| WTT HR Expenditure, B | 0.8 |
| WTT Cardiac Effort, B/m | 0.72 |
| WTT Pre-Walk Dyspnea | 0.61 |
| WTT Post-Walk Dyspnea | 0.7 |
| WTT Pre-Walk Tiredness | 0.62 |
| WTT Post-Walk Tiredness | 0.72 |

WTT = Walk.Talk.Track, ICC = Intraclass Correlation, NYHA FC = New York Heart Association Functional Class, MAD = Mean Amplitude Deviation, HR = Heart Rate, B = beats

**Table E3. Description of the extracted Digital Measures**

| WTT Variable | Unit | Description |
|---|---|---|
| *WTT 6-Minute Walk Distance Raw ($6MWD_R$)* | m | Total distance estimated based on Apple's proprietary algorithms. |
| *WTT 6-Minute Walk Distance Calibrated ($6MWD_C$)* | m | Total distance estimated from a mixed effects linear regression model with a random intercept to account for repeated measures within a participant. Fixed effects included age, sex, weight, height, and number of steps taken. The model was fit on pairs of concurrent gold standard indoor 6-minute walk distance ($6MWD_{ATS}$) and WTT walk data. Subsequently, model performance was evaluated on gold-standard outdoor 6MWTs conducted on the same day. |
| *WTT Steps* | count | Total number of steps during the 6MWT based on Apple's proprietary algorithms. |
| *WTT Total Mean Amplitude Deviation (MAD)* | g | The numerical integral (using the Trapezoidal rule) of the mean amplitude deviation (3). This is a measure of physical activity intensity based on raw accelerometer readings. Before integrating, the gravity component was removed from each accelerometer axis and the signal was resampled to 15s epochs for which the MAD was calculated. |
| *WTT HR Reserve* | bpm | The heart rate reserve (HRR) was calculated by subtracting resting heart rate from the estimated maximum heart rate of a participant, where $HR_{max} = 220 - age$ and $HR_{rest} = Q^{05}(HR_{day})$. The last term refers to the 5th quantile of a participant's heart rate reading taken on the day the walk was performed. |
| *WTT Chronotropic Index* | unitless | Estimated by the formula below, where $HR_{peak}$ refers to the WTT Peak HR measure. The HRR is linked to the same day the walk was performed. $$\frac{HR_{peak} - HR_{rest}}{HRR}$$ |
| WTT HR Recovery *(HRR@1)* | bpm | Refers to the estimated heart rate recovery 1 min post walk. This was estimated by taking the last HR sample of the one-minute resting period after the walk. |

| | | |
|---|---|---|
| *WTT Peak HR* | bpm | Defined as $Q^{95}(HR_{walk})$, the 95th quantile of heart rate samples during the walk. |
| *WTT HR Expenditure (HRE)* | beats | Following Lachant et al., this refers to the total number of heartbeats throughout the walk (4). To obtain HRE, the Apple-provided HR signal was resampled to 15s bins and subsequently numerically integrated using the trapezoidal rule. |
| *WTT Cardiac Effort (CE)* | beats/m | Refers to a measure of cardiovascular effort, hypothesized to capture right ventricular function in PAH patients (5). CE is the ratio between HRE and 6MWD (where not reported otherwise we used WTT $6MWD_C$). |
| *WTT Pre-Walk Dyspnea Rating* | scale | Refers to the answer participants reported just before performing a WTT walk. To gauge dyspnea participants were asked the question, "How short of breath are you now?". They were given the following options – where the number in brackets refers to the numeric rating: (0) "Nothing at all."; (1) "Slight."; (2) "Moderate."; (3) "Severe."; (4) "Very severe.". |
| *WTT Post-Walk Dyspnea Rating* | scale | Refers to the same as the WTT Pre-Walk Borg Dyspnea Index but asked just after the WTT walk. |
| *WTT Pre-Walk Tiredness Rating* | scale | Refers to the answer participants reported just before performing a WTT walk. To gauge tiredness participants were asked the question, "How tired are you now?". They were given the following options – where the number in brackets refers to the numeric rating: (0) "Nothing at all."; (1) "Slight."; (2) "Moderate."; (3) "Severe."; (4) "Very severe.". |
| *WTT Post-Walk Tiredness Rating* | scale | Refers to the same as the WTT Pre-Walk Dyspnea Rating but asked just after the WTT walk. |

### Supplementary Figures

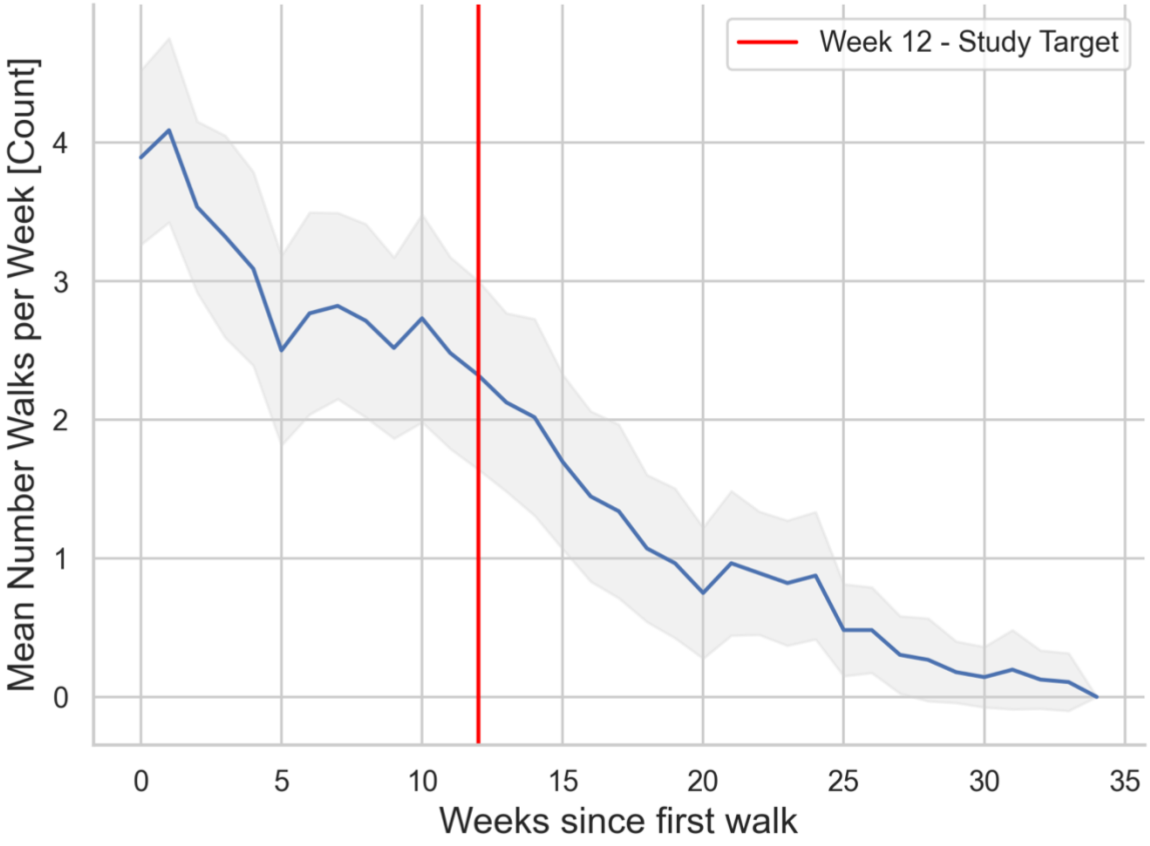


**Figure E1. Average number of app derived walks per week across all participants.** The red line denotes the end of the official 12-week study target duration, while the grey area refers to the 95% confidence interval.

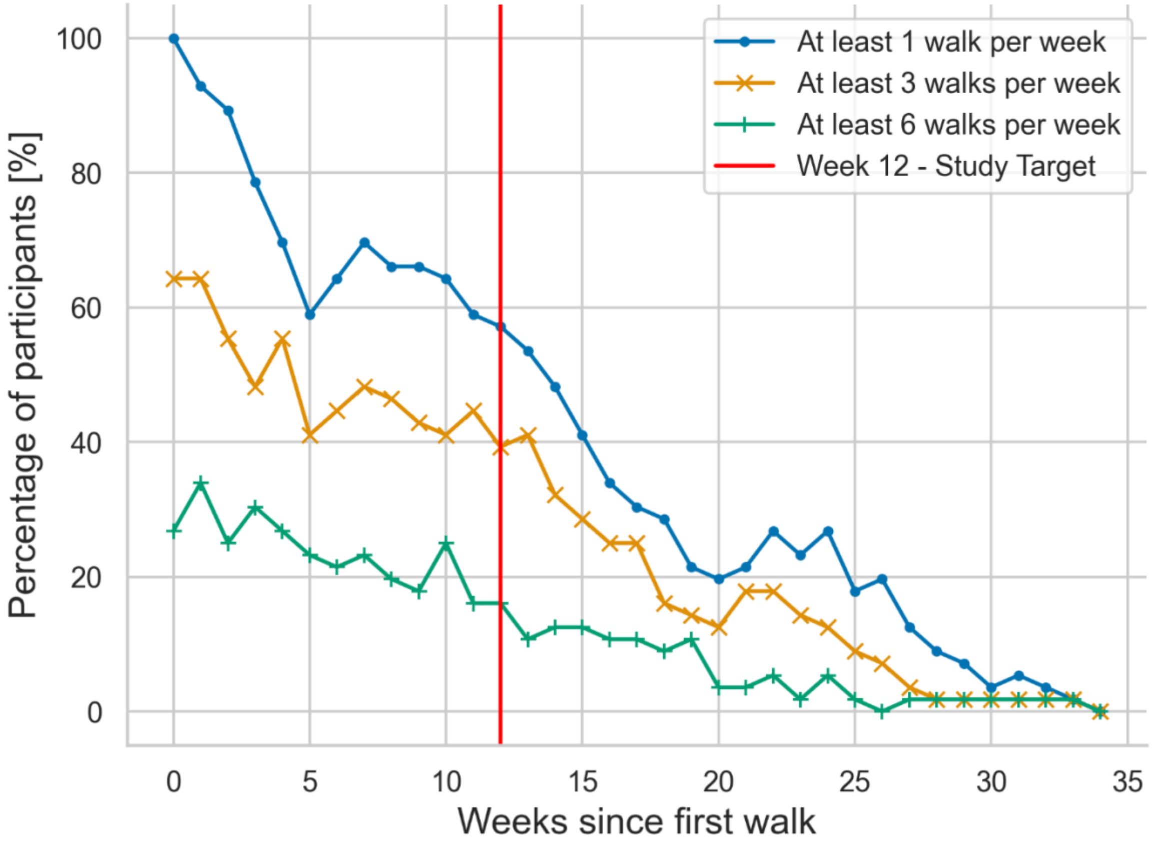


**Figure E2. Percentage of participants that performed at least 1, 3, or 6 walks per week over the study duration (red line) and beyond.**

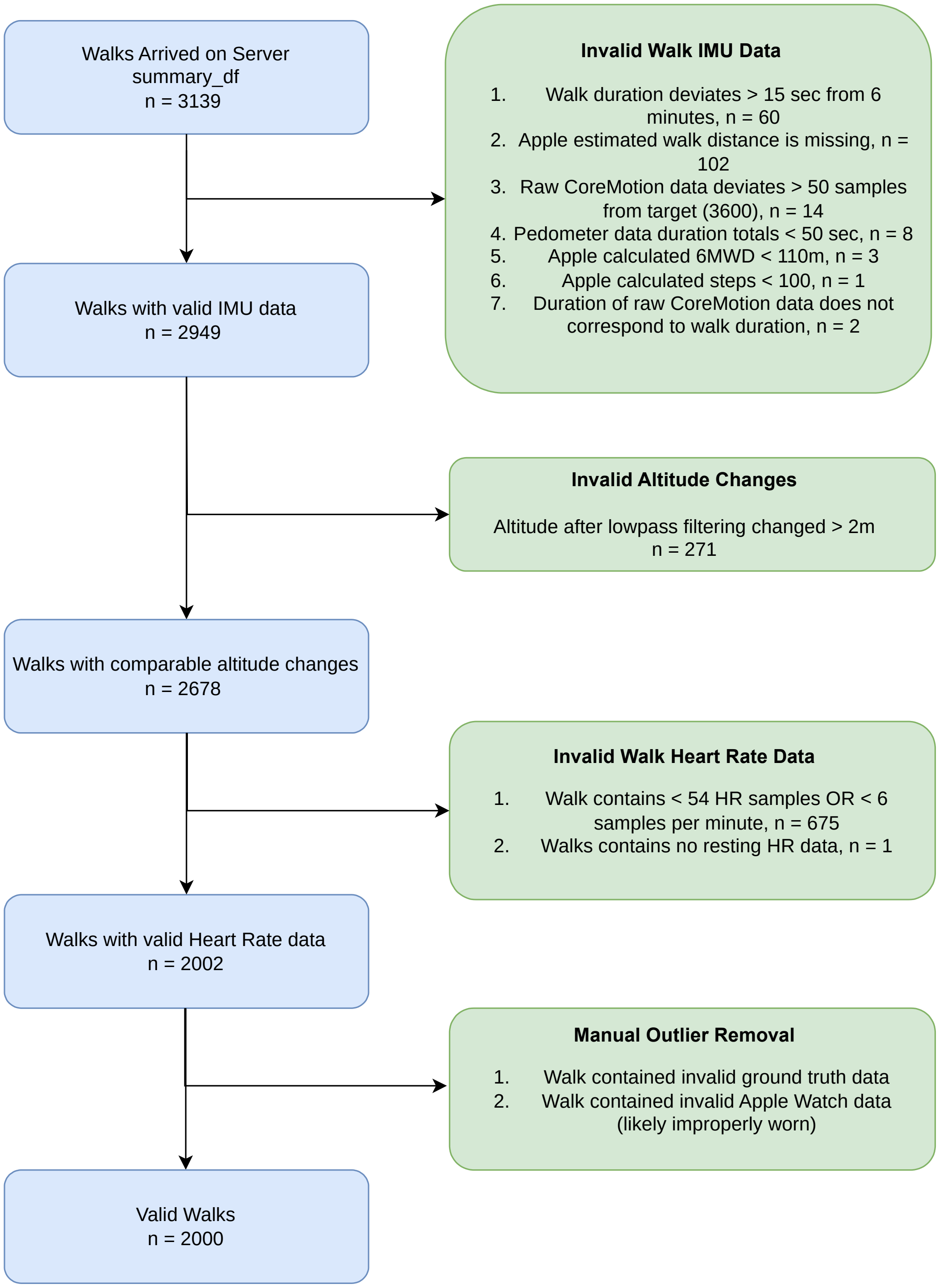


**Figure E3. A flowchart diagram of the filtering process to remove app-derived walks with invalid data.**

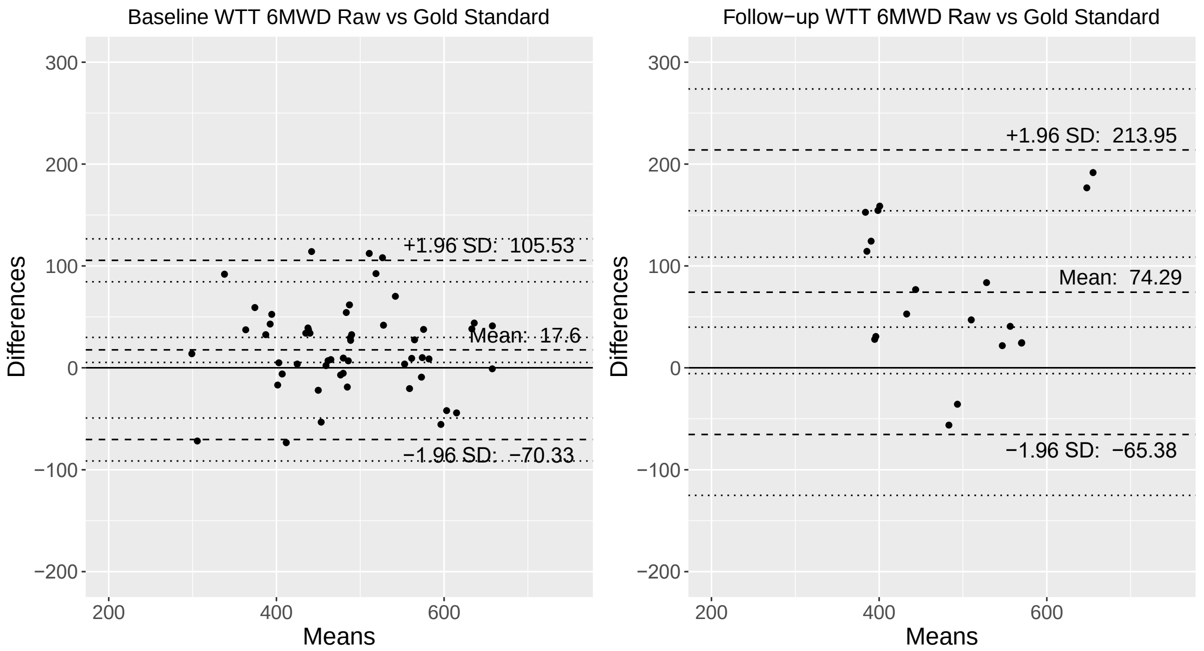


**Figure E4. Baseline vs follow-up agreement of WTT 6MWD and In-Clinic 6MWD.** Shows agreement between the average at-home WTT 6MWD Raw values within 2 weeks of the clinic visit and the gold standard 6MWD values at the clinic visit. The left figure shows the adjacent at-home values of the baseline clinic visit, while the right shows the same for the 3-month follow-up visit. The agreement seems markedly lower for the follow-up, when compared to the baseline, both in terms of bias and variation. In both scenarios, the at-home values underestimate the gold-standard 6MWD.

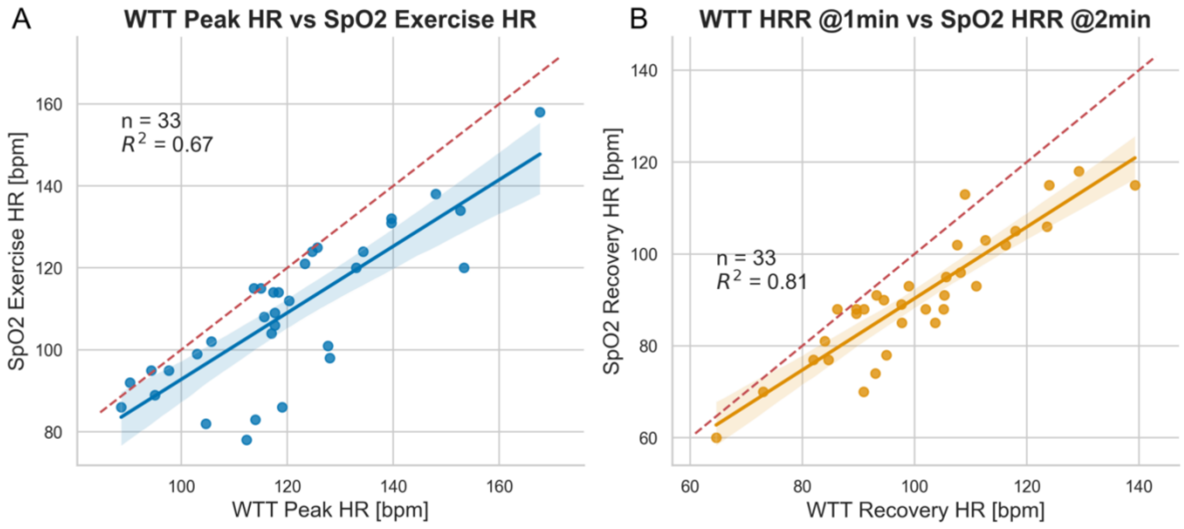


**Figure E5. Apple Watch derived WTT Peak HR and WTT Heart Rate Recovery against their in-clinic oximeter monitor based versions. (A)** Shows in-clinic oximeter monitor derived heart rate, measured just after the walk, against the peak heart rate recorded by the WTT app using the Apple Watch. **(B)** displays heart rate after one minute of recovery (post walk) for the WTT app and 2 minutes after the walk measured by the oximeter. All presented data stems from in-clinic walks.

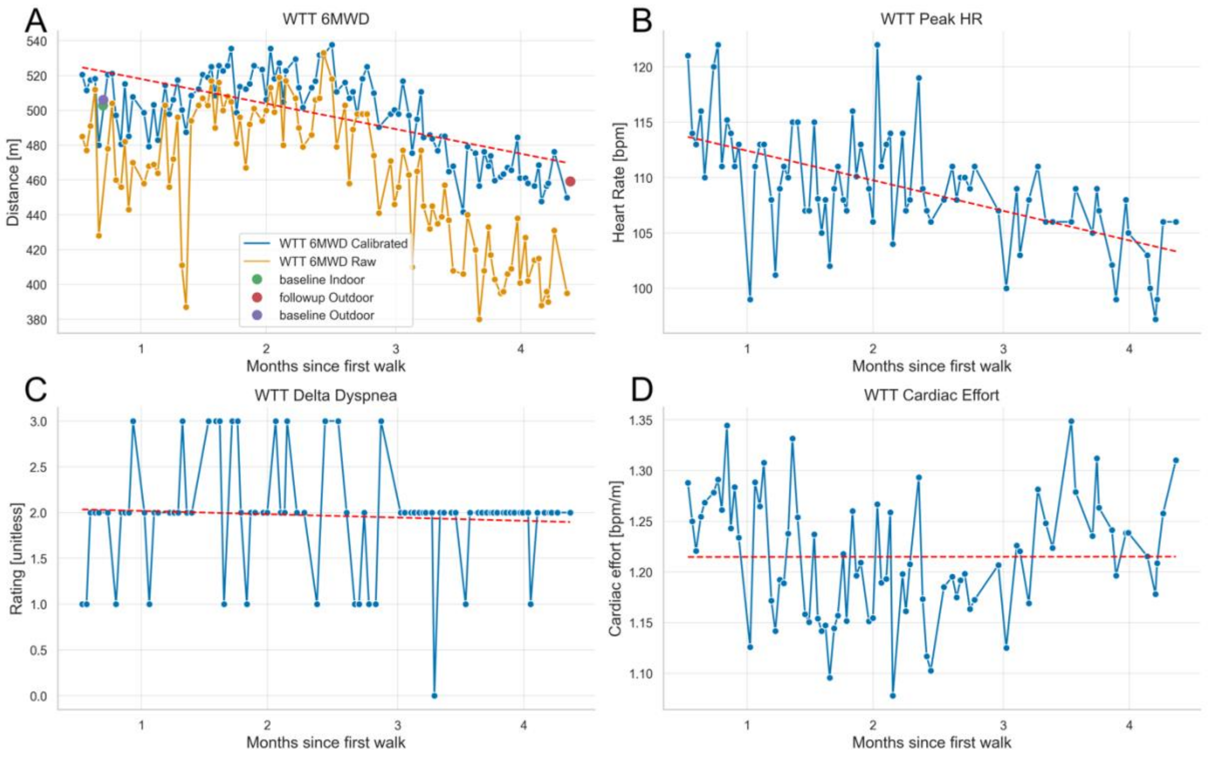


**Figure E6. An example case of a participant with longitudinal data matching the observations in the clinic. (A)** shows the raw and calibrated app-derived walk distances. In-clinic baseline and follow-up 6-minute walk distances are marked. The measured app-derived walk distances recorded in the community setting match the in-clinic measurements quite well. **(B)** details app derived peak walk heart rate over time, which showed a relatively steady decline. **(C)** displays the difference in perceived dyspnea ratings before and after a walk, it is apparent how no clear trend emerges. **(D)** shows an attempt to capture effort through the notion of cardiac effort (the number of heart beats divided by the walked distance). Again, with no clear trend present.

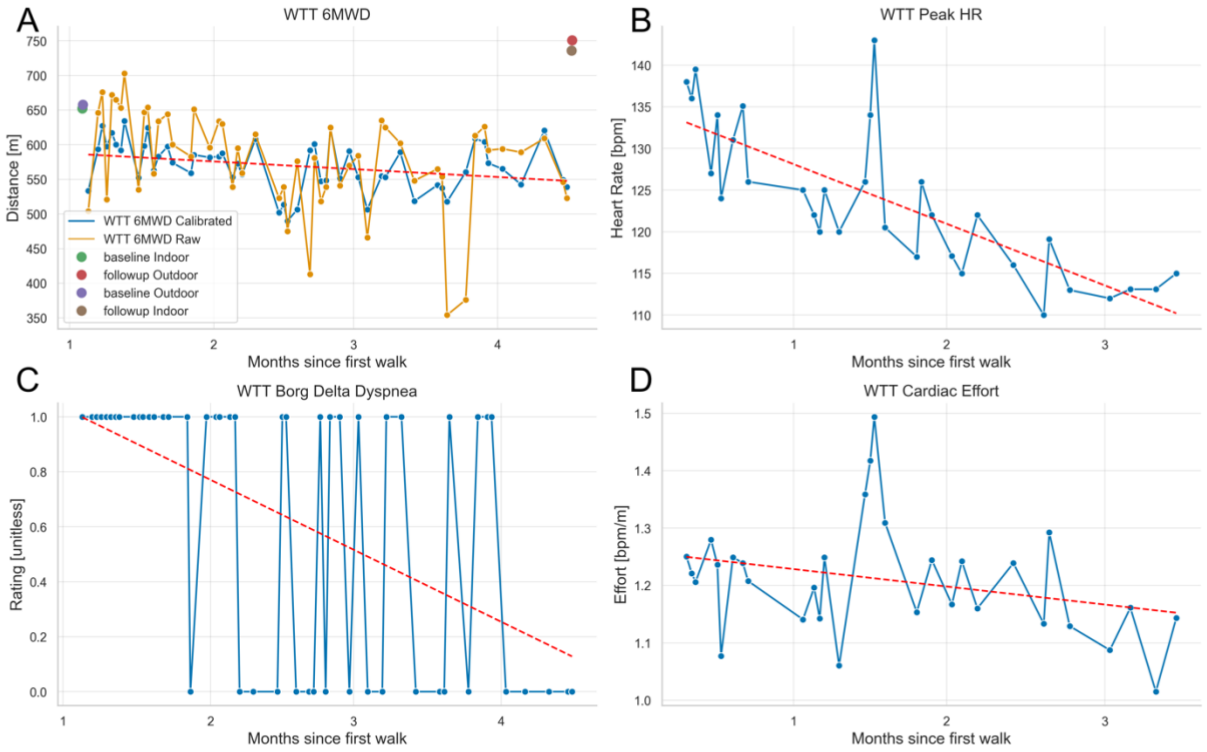


**Figure E7. An example case of a participant with longitudinal data diverging from the observations made in the clinic. (A)** shows the raw and calibrated app derived walk distances. In-clinic baseline and follow-up 6-minute walk distances are marked. The app derived walk distances do somewhat match the in-clinic measurements at the beginning but seem to diverge over time, leading to a larger discrepancy with the follow-up in-clinic measurements. **(B)** details app derived peak walk heart rate over time, which show a steady decline. **(C)** displays the difference in perceived dyspnea ratings before and after a walk, a clear trend emerges where walks seem to lead to less perceived dyspnea over time. **(D)** shows an attempt to capture effort through the notion of cardiac effort (the number of heart beats divided by the walked distance). There seems to be a slight decrease.

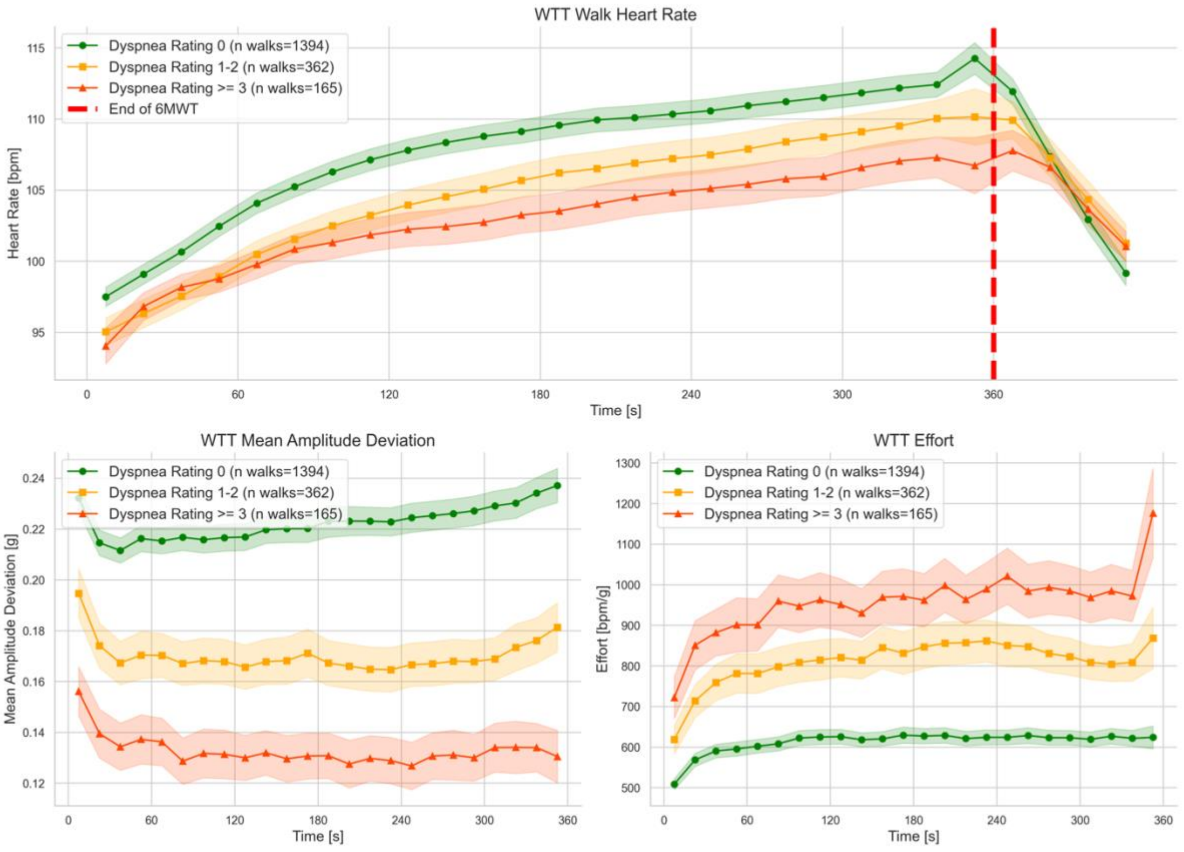


**Figure E8. Continuous WTT 6MWT Variables Grouped by Self-Rated Dyspnea Ratings.** Multiple time binned WTT variables across different patient-reported post-walk Dyspnea Index ratings: how short of breath are you: 0=nothing at all; 1=slight; 2=moderate; 3=severe; 4=very severe. This highlights how, on average, walks with a specific post-walk dyspnea rating evolved over the 6-minute walk duration before the rating. A) WTT Heart Rate refers to the Apple provided heart rate throughout and 1min after the walk; B) WTT Mean Amplitude Deviation is a measure of physical activity intensity based on raw accelerometer data throughout the walk; C) WTT

Effort relates to continuous pulse estimates to continuous physical using a ratio, where higher values indicate greater effort.

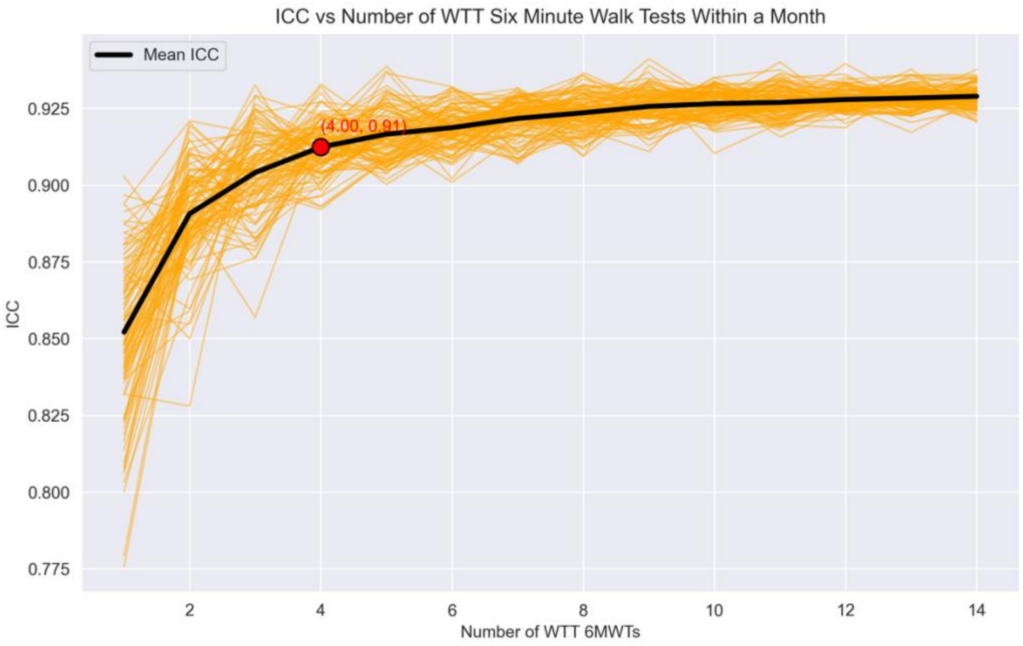


**Figure E9. Intraclass correlation (ICC) values for different numbers of walks per month and participants.** Highlighted is the optimal tradeoff between the highest ICC and the smallest number of walks per 4-week interval (month) in red. 100 simulations are drawn and plotted in orange, as well as the mean of those, plotted in black.

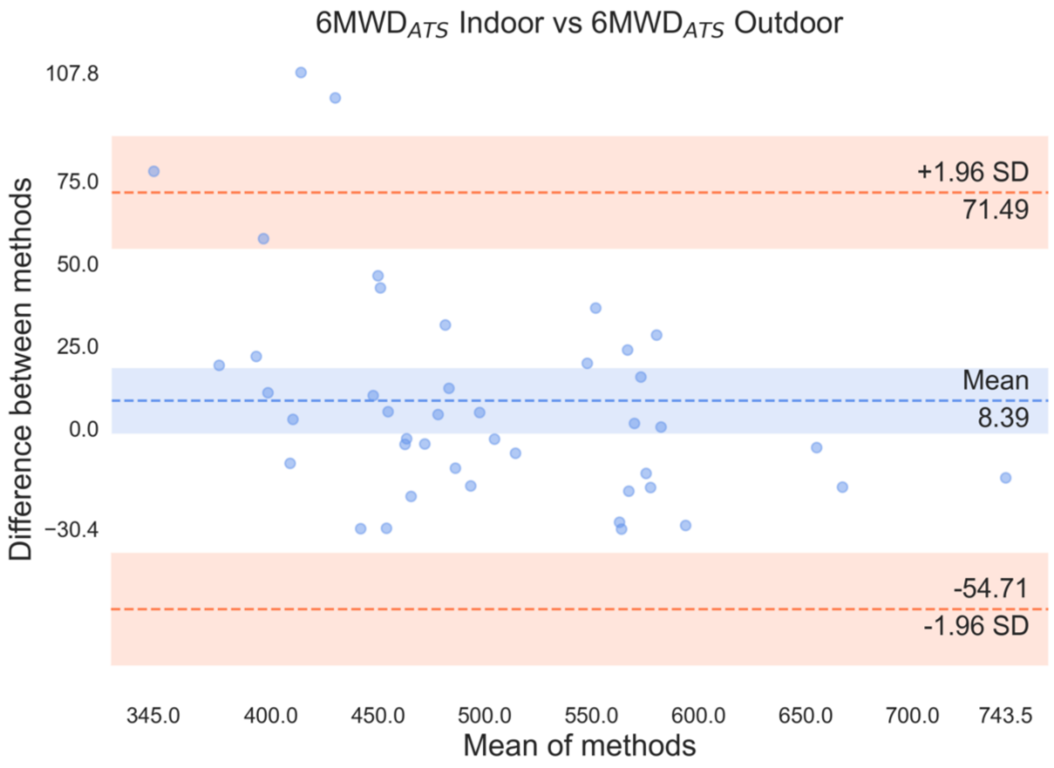


**Figure E10. Bland Altman Analysis of Same-Day Gold Standard 6MWDs.**

Shown is the agreement between the same-day pairs of in-clinic conducted and rated 6MWTs that were performed during the study. One 6MWT was conducted indoors and one outdoors but both were conducted under clinical supervision and within a few hours from each other.

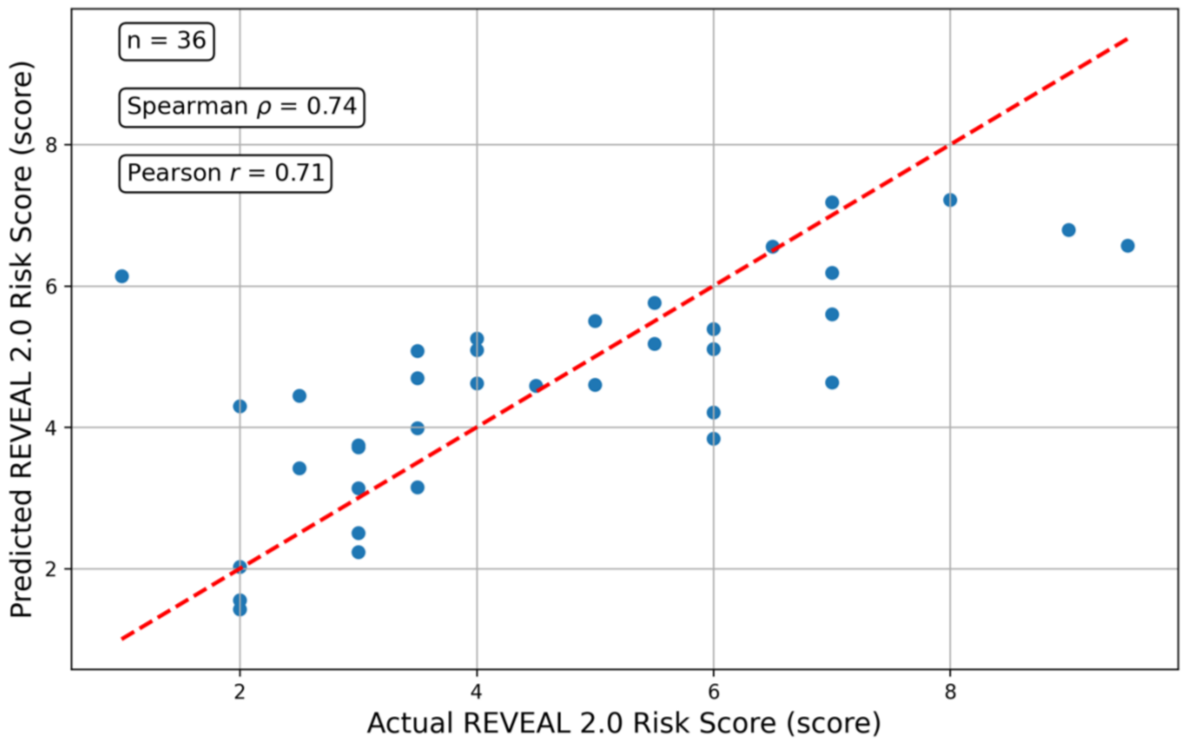


**Figure E11. Actual REVEAL 2.0 vs Predicted REVEAL 2.0 Risk Scores.**

Shows multivariate linear regression predictions of REVEAL 2.0 risk scores based on the 10-dimensional latent representations extracted by the used deep learning/ ordinary differential equation heart rated response modelling approach. The red line represents a perfect fit. Spearman and Pearson correlation coefficients as well as number of participants (=data points) are displayed as annotations.
